# Estimation of the Reproduction Number for COVID-19 Based on Latest Vaccination Results and the Timing for Herd-Immunity: Prospect for 2021

**DOI:** 10.1101/2021.03.25.21254362

**Authors:** Steven Suan Zhu, Enahoro Iboi

## Abstract

This study examined four countries Israel, United States, United Kingdom, and Serbia and present their possible vaccination trajectories into 2021. We found that populations in all the four countries are relaxing and taking the advantage of the benefit of an increasingly immunized community hence, experiencing a rising phase of ℛ _*c*_(*t*). The United States is of particular concern, due to its fast rising ℛ_*c*_(*t*) in comparison to other countries, potentially generating another wave of infection. Due to aggressive vaccination program, continued implementation of restrictive measures, or both, in all countries we analyzed, present a cautiously optimistic outlook at controlling the pandemic toward the latter part of 2021. We also found that despite a significant fraction of the population in selected countries being immunized, no countries other than Israel has its ℛ_*c*_(*t*) reached its intrinsic ℛ_0_ value. Based on our proposed methodology for deriving ℛ_0_, our prediction shows that Israel’s indigenous COVID-19 daily ℛ_0_ is approximately 2.2 based on its latest data.

## 1 Introduction

When COVID-19 Pandemic Struck the world in late December 2019, organizations, companies, and governments raced for the development of vaccine against the disease. Throughout 2020, more than a few dozens of vaccines were under development. Toward the fall of 2020, several vaccines entered the late phase of clinical trial III. [1] Then, Russia became the first nation to put its self-developed *Sputnik V* vaccine into usage domestically [2]. On December 2, 2020, the United Kingdom (UK) gave regulatory approval for Pfizer–BioNTech vaccine, [3, 4] becoming the first country in the Western world to approve the use of any COVID-19 vaccine on humans. By end of 2020, many countries including the United States (US) and the European Union (EU) have authorized or approved the Pfizer-BioNTech vaccine[5]. Bahrain and the United Arab Emirates (UAE) granted emergency marketing authorization for *BBIBP-CorV*, manufactured by Sinopharm from China [6, 7]. Moderna which is the first RNA vaccine was approved in the US in late December 2020 [8]. By the beginning of 2021, a significant number of countries around the world has at least implemented some vaccination delivery program to its citizens. Among them, several countries such as Israel, Seychelles, UAE, US, UK, Chile, and Serbia stood out from the rest with rapid and ambitious vaccination strategies.

The objective of this study is to use the rich available vaccination data accumulated during the early months of 2021 from the aforementioned countries to gain better understanding of the effectiveness of the vaccines at curbing the pandemic and human behavioral changes in response to the vaccine. Israel was selected since it lead others in terms of vaccination administration on its citizens. US was also selected due to its absolute population and infection size, UK and Serbia are included since they are the most aggressive countries in terms of its vaccination program in Europe. We will put our theoretical model to test with real world data, and make predictions about the progression of the pandemics in the presence of vaccinations, the timing of herd immunity, and ultimately estimate the ℛ_0_ value of COVID-19 infection. Many mathematical models have been proposed for modeling COVID-19 and its subsequent vaccination program (see for instance [9, 10, 11, 12, 13, 14, 15, 16, 17, 18, 19, 20]). In general, epidemic models broadly explores the evolution of disease processes over time, and explore patterns and correlations in data using regressions and time series analysis. Epidemic models are divided into deterministic, stochastic, network, and agent based. Models described as continuous in time uses differential equations (compartmental models), while discrete-time models uses the difference equations [19, 18, 20].

In our study, we are adopting a hybrid modeling approach, involving dynamic and statistical modeling. We use statistical regression to derive the time dependent ℛ_*c*_ (*t*), which is the ℛ_0_ value change in response to the implementation of control measures to account for the gradual refinement in, or improvement of, these control measures. We found, based on earlier works with fairly good approximation, despite the non-autonomous characteristics of time dependence of parameters such as ℛ_*c*_ related to the implementation of control measures [21], can be worked around without devolving into the intricate inner working interactions by the simple assumption, that a momentum is gained so that the cooperation and change of behaviors between the citizens and the governments is maintained once quarantine isolation measures takes in place [22]. As a result, a measure tends to be enforced in place steadily for a long period of time rather than dynamically changes on a weekly or even daily basis [22]. In a sense, it is the same assumption used in economic theory, that one can assume the participants are rational players act cautiously based on their assessment of the situation at the time. Therefore, the trend observed from the extrapolation from historical time series can be applied for future predictions. We adopt a deterministic discrete-time model using difference equations in units of day to predict the epidemic progression of each country through simulation. No stochastic model is necessary since all countries under study has a population size of at least 7 million.

In our previous papers [23, 24, 25, 26, 27], we have outlined this deterministic model for viral infection and its instantiation for COVID-19 in particular. The final infection ratio within any given population without any artificial intervention and have shown that the final infection ratio in a natural, non-interfered pandemic must fall within a specified lower and upper bound given by 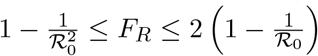, where ℛ_0_ is the intrinsic ℛ_0_ value of the disease. We also outlined that all pandemic progression can be summarized within 4 possible scenarios:

1. fixed ℛ_0_ + fixed cumulative vaccination *V* (*t*), minimal lockdown measures and no vaccinations,
2. variable ℛ_*c*_(*t*) + fixed cumulative vaccination *V* (*t*), lockdown measures with no vaccinations,
3. fixed ℛ_0_ + variable cumulative vaccination *V* (*t*), minimal lockdown measures but with constant vaccination rates
4. variable ℛ_*c*_(*t*) + variable cumulative vaccination *V* (*t*), lockdown measures and constant vaccination rates.

This study focus on the last scenario since we found that in all countries we analyzed, the pandemic falls under the last scenario, that is, variable ℛ _*c*_(*t*) + variable cumulative vaccination *V* (*t*).

The remainder of the work in this paper is organized as follows. Section 2 outlines the statistical method for deriving ℛ_*c*_ (*t*) and the construction of deterministic discrete-time model using difference equations in units of day, and the statistical derivation of ℛ_0_ based on known ℛ_*c*_ (*t*), susceptible ratio *S* (*t*), and daily observed infection *I*^*d*^ (*t*). Section 3 outlines pandemic projections for Israel, the US, UK, and Serbia. Section 4 and 5 outlines discussions and conclusions regarding the model.

## 2 Materials and Methods

### 2.1 Derivation of the control ℛ_*c*_ (*t*) and the construction of deterministic, recursive, discrete-time simulation model

The immunity ratio within the entire region of interest will now be derived. Let *N* be the total population of our region of interest and *I* (*t*) be the cumulative number of infections observed on day *t*. Let *V*_1_ (*t*− *t*_*ϵ*_) be the cumulative population that received at least one dose by day *t* and *V*_2_ (*t* − *t*_*ϵ*_) be the cumulative population that received 2 doses, where *t* is the current day. The parameter *ϵ* represent the delay of the onset of antibodies after immunization, so that the effective immunized population at the current day observed always correlate with cumulative immunized population size at an earlier date. Let *e*_1_ be the efficacy for inducing antibodies based immunity with 1 dosage, and *e*_2_ is the efficacy for inducing antibodies based immunity with 2 dosages so that

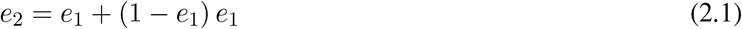

The immunity ratio *E* (*t*) within the entire population can be computed based on the following equation:

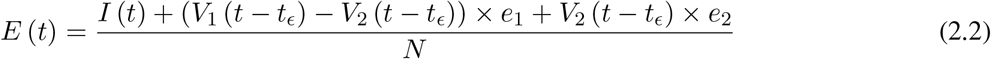

We denote 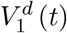 to be the daily vaccination number, which is the difference between the cumulative number of vaccinations on any consecutive days. Hence,

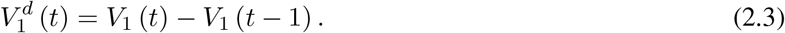

Similarly, let *I*^*d*^ (*t*) represent the daily new infections, then

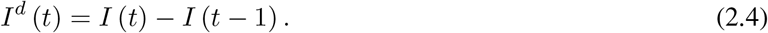

The control reproduction number ℛ_*c*_(*t*) which is defined as the daily rate of change in the number of new infections observed divided by the ratio of those who remains susceptible is given by

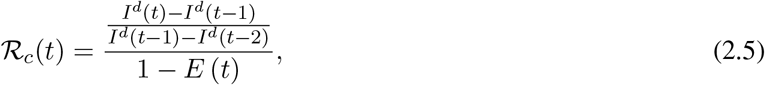

Using the 7 days moving average 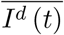 instead of 1 day to derive the daily rate of change to smooth the cyclic fluctuations observed in COVID-19 cases,[28] equation 2.5 becomes

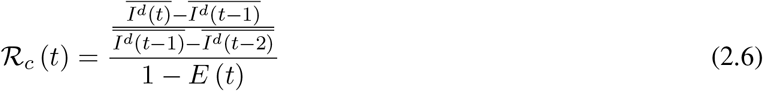

Based on the historical time series of ℛ_*c*_(*t*), which can be expressed as:

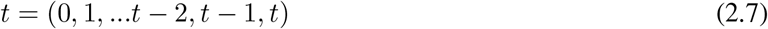

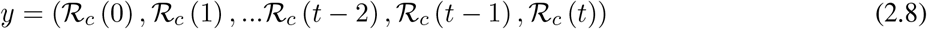

we find the best fit (polynomial or linear fit) that best extrapolate future trends as:

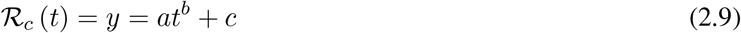

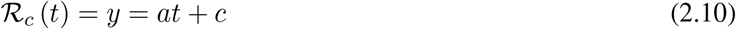

which generalize ℛ_*c*_(*t*) into a closed form by incorporating the past and projecting into the future.

The simulation will be carried out using the following recursive equation:

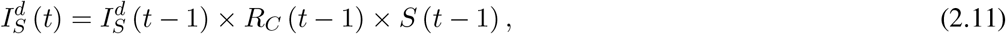

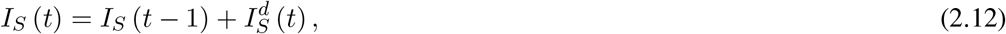

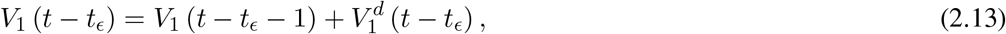

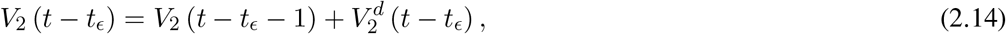

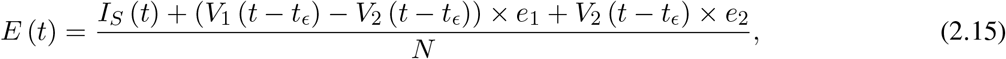

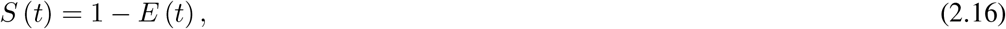

where the daily infection 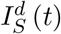 is first computed for the simulated results based on previous day’s infection and the latest change on control ℛ_*c*_(*t* − 1) as well as the remaining number of susceptible. Then the cumulative number of infected *I*_*S*_(*t*) with 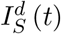 are updated. Next, the cumulative vaccination number *V*_1_ (*t* − *t*_*ϵ*_) and *V*_2_ (*t* − *t*_*ϵ*_) are updated with daily vaccination increase. Further, the current effective immunization ratio *E*(*t*) is updated. Finally, the remaining susceptible ratio is obtained from *E*(*t*).

### 2.2 Derivation of Intrinsic ℛ_0_ from ℛ_*c*_(*t*), *S*(*t*), and daily infection *I*^*d*^(*t*)

For a disease such as COVID-19, we do not know the upper bound of ℛ_0_ and there is no guarantee that it can be derived even when 100% immunity is achieved. To see why, we define the first day when vaccinations are introduced as *t*_*F*_ and the last day when vaccinations are completed as *t*_*L*_, so that all days of *t* falls within *t*_*F*_ *≤ t ≤ t*_*L*_, then imagining that dosages are administered at a certain rate *V* ^*d*^(*t*), infection are introduced at a certain rate *I*^*d*^(*t*), and the control ℛ_*c*_(*t*) is also rising at a certain rate. So the controlled endemic equilibrium is:

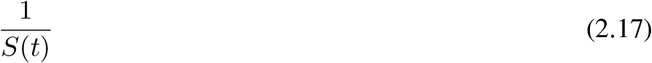

That is, the inverse of the remaining susceptible population ratio, and 0 *≤S*(*t*) ≤1. The remaining susceptible population ratio is expressed as:

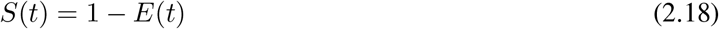

When the controlled endemic equilibrium brought by 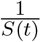 exceeds the ℛ_*c*_(*t*) rate at all times *t*_*F*_*≤ t≤ t*_*L*_, the entire population becomes immunized before its behavior returned to complete normalcy, so that:

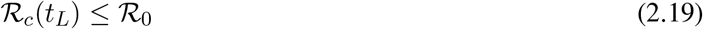

In the most extreme case, all vaccinations are delivered instantly to all populations while the entire population was under a complete lockdown with a daily control ℛ_*c*_ *<* 1 as:

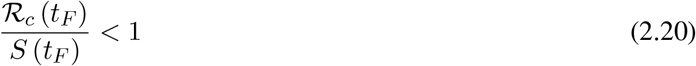

When the society returned to normalcy, no additional daily ℛ_*c*_ can be observed since no additional cases are reported. In fact, no additional cases can be observed even before *t*_*L*_ is reached. Therefore, the final derived ℛ_0_ *<* 1 based on ℛ_*c*_. Nevertheless, in general, full vaccinations are achieved after a period of time. During this time, control ℛ_*c*_ steadily rises but yet to reach its intrinsic value ℛ_0_. As a result, at the end of vaccination, one can only predict that ℛ_0_ ≥ ℛ_*c*_. When vaccination program lasts long enough so that there is enough time for ℛ_*c*_ (*t*) reaching its intrinsic ceiling value of ℛ_0_. Then it can be observed based on the discrepancy between the projected path and the actual path. We define that there exists a time *t*_0_ in which *t*_*F*_ *< t*_0_*< t*_*L*_, so that any time *t* occurs later than *t*_0_ as *t* > *t*_0_, we have:

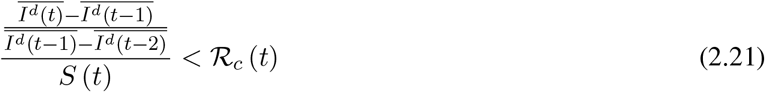

That is, the observed daily infection ℛ_*C*_ values starts to fall below the expected continually rising ℛ_*c*_ (*t*) curve. In other words, ℛ_*c*_ (*t*) is no longer increasing and the intrinsic ℛ_0_ value is reached. We can also detect such change by:

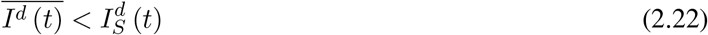

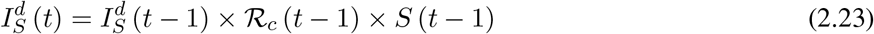

That is, the number of reported infections starts to fall below the number predicted for a continually rising ℛ_*c*_ (*t*) curve. Where 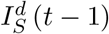 is the recursively simulated/expected infection numbers on day *t* − 1, the product with ℛ_*c*_ (*t* − 1) × *S* (*t* − 1) yields the recursively simulated/expected infection numbers on day *t* as 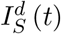 The earliest date in which the intrinsic ℛ_0_ can be derived is then *t* = *t*_0_:

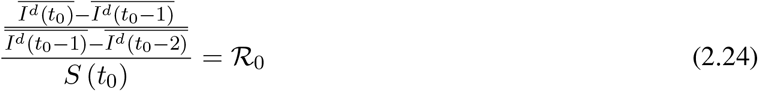

But one should able to derive ℛ_0_ for any *t* > *t*_0_ and certainly we have:

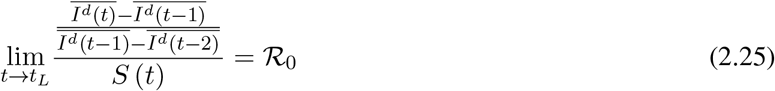

Figure 2.1 shows a model simulation, with a ℛ_*c*_ (*t*) = 0.0002*t*^_2_^ + 1, and an intrinsic ℛ_0_ = 2 and initial seed infection size = 40 and total population of 10000, both graph shows the endemic equilibrium required by ℛ_*c*_(*t*) and the remaining susceptible ratio *S*(*t*) at any given time. In the top graph, vaccination at a rate of 90 daily is fast enough so that the last infection is observed well before ℛ_*c*_(*t*) returned to its intrinsic endemic equilibrium 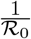. As a result, the remaining susceptible ratio is reduced solely by vaccination and true ℛ_0_ can not be obtained due to an absence of observable infection. In the bottom graph, vaccination at a rate of 5 daily is slow enough so that the last infection occurs well after ℛ_*c*_ (*t*) returned to its intrinsic endemic equilibrium 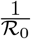. As a result, there exists an observation window to confirm the true value of ℛ_0_.

**Figure 2.1:**
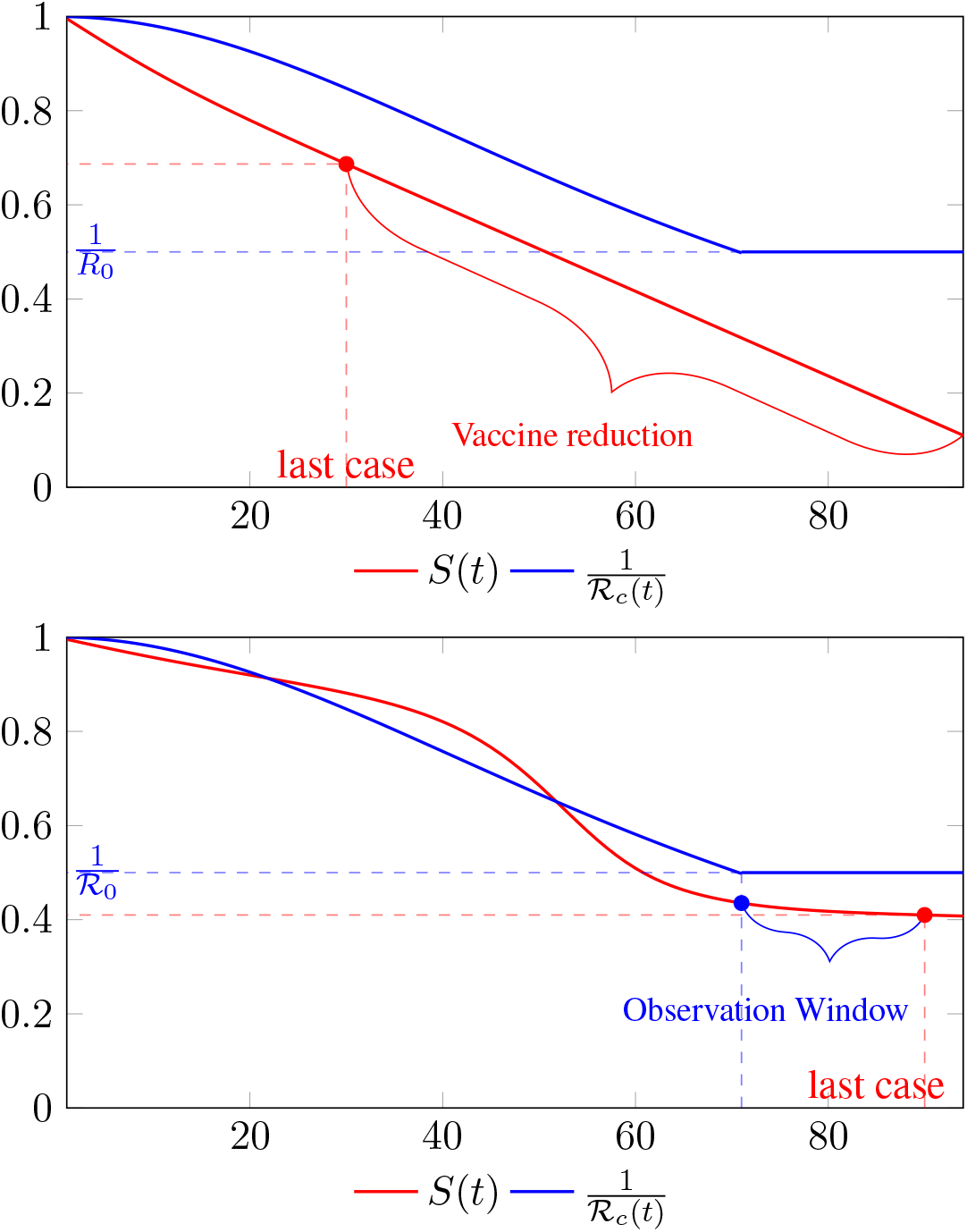

Figure 2.2 shows a model simulation, with a ℛ_*c*_ (*t*) = 0.00077*t*_2_ + 1, and an intrinsic ℛ_0_ = 2 and initial seed size =100, and from left to right: *V* ^*d*^ (*t*) = 400, *V* ^*d*^ (*t*) = 200, *V* ^*d*^ (*t*) = 150, *V* ^*d*^ (*t*) = 110, and *V* ^*d*^ (*t*) = 4, so each day 400, 200, 150, 110, and 4 people vaccinated so immediately removed from remaining susceptible respectively. the final daily infection number 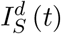 is displayed by calendar days. The gray line indicates that given enough time *t* ≥ *t*_0_ = 37, the rising ℛ_*c*_ (*t*) has returned the control ℛ_*c*_ back to ℛ_0_. Only under *V* ^*d*^ (*t*) = 110, and *V* ^*d*^ (*t*) = 4, vaccination scheme was slow enough so that _0_ can be effectively derived based on non-zero values of infections. Under cases *V* ^*d*^ (*t*) = 400, *V* ^*d*^ (*t*) = 200, and *V* ^*d*^ (*t*) = 150, zero infections are reported before the rising ℛ_*c*_(*t*) has returned the control ℛ_*c*_ back to ℛ_0_. Therefore, aggressive vaccination results in last infection occurs before ℛ_*c*_(*t*) returned to its ℛ_0_ value. Since ℛ_*c*_ values can be derived only based on daily infection changes, once zero infection is reached, there is no way to know the true ℛ_0_ of the disease. Under those scenarios, ℛ_*c*_ (*t*_*L*_) *<* ℛ_0_.

**Figure 2.2:**
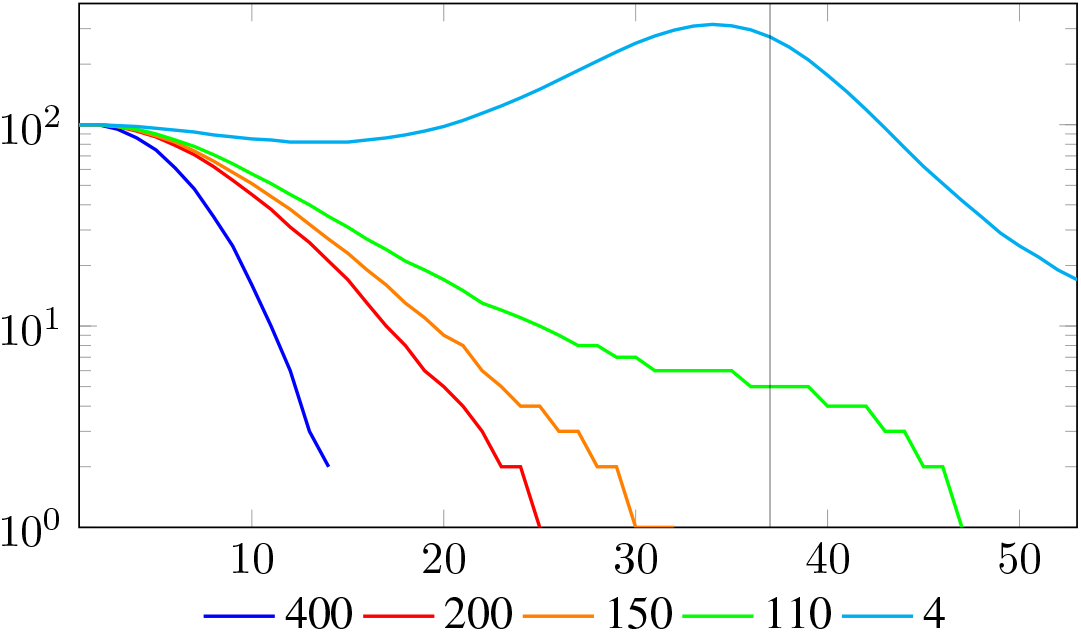
In this model simulation with aggressive vaccination strategy > 110 vaccinations daily, the last infection is observed before day 36, when ℛ_*C*_ (*t*) returned to its ℛ_*c*_ (*t*_*L*_)

Figure 2.3 shows the final daily infection number 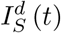 based on *ν*_*c*_ (*t*) curves, one is capped at intrinsic ℛ_0_ = 2 and the other is capped at intrinsic ℛ_0_ = 10, the one capped at higher ℛ_0_ experienced 2 waves of infection as rising ℛ_*c*_ (*t*) repeatedly exceeds 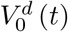. We also show that there exists a *t*_0_ = 38, for which *t*_*F*_ *< t*_0_ *< t*_*L*_, and 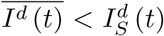 in which we assume that the ℛ_*c*_(*t*) curve capped at ℛ_0_ = 2 represents 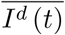, and the other is capped at intrinsic ℛ_0_ = 10 represents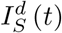, despite the fact that both are 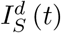.

**Figure 2.3:**
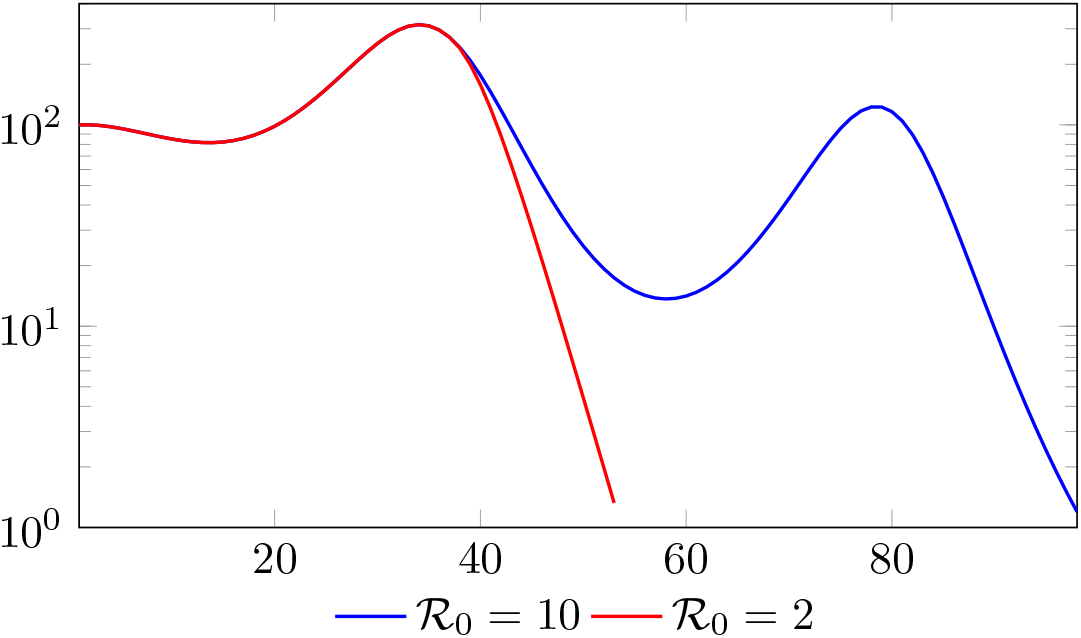
In this model simulation, 2 waves of infections is observed for ℛ_0_ = 10 compared to only 1 for ℛ_0_ = 0) final daily infection number 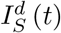 based on *v*_*c*_ (*t*) curves with *R*_0_ = 2 and *R*_0_ = 10

Of course, our assumption is based on the fact that once lockdown measures are implemented in place, its removal is always smooth and gradually in reality, corresponding to smooth, steadily rising *n*th degree polynomial without any discontinuities. In fact, a steadily rising *n*th degree polynomial ℛ_*c*_(*t*) can render ≥ 1 waves of infections as it returns to complete normalcy if ℛ_*c*_(*t*) exceeds the vaccination rate *V* ^*d*^(*t*) at all times.

## 3 Results

### 3.1 Israel Projections

Israel uses Pfizer vaccine, with vaccine efficacy after the first dose to be *e*_1_ = 0.76 and *e*_2_ = 0.9464 to be the efficacy after the second dose, total population *N* = 9053000, and *I*(*t*) = 796465 [29, 30, 31]. We obtain both time series for *V*_1_ (*t* − *t*_*ϵ*_) and *V*_2_ (*t* − *t*_*ϵ*_), where *t* is March 5, 2021, and *t*_*ϵ*_ = 8 [32]. Hence, the immunity ratio curve for Israel is depicted in Figure 3.1. It is worth mentioning that the current weighted immunity achieved for Israel have just exceeded 50%.

**Figure 3.1:**
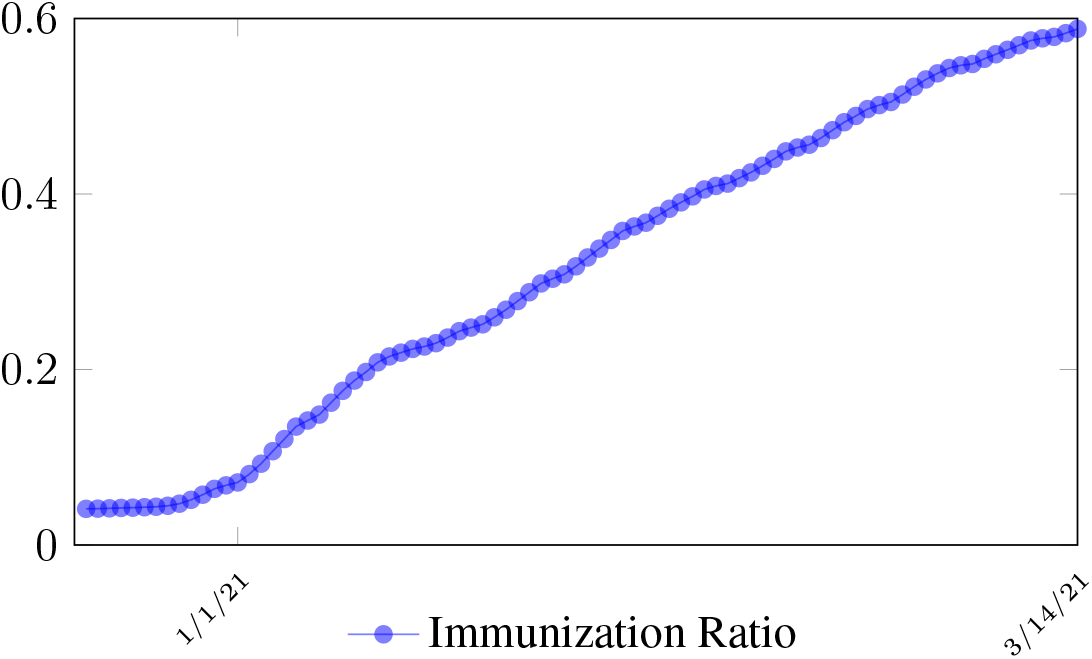
Israel effective immunization ratio

Figure 3.2 shows the final control reproduction number ℛ_*c*_(*t*) curve is a rising one, indicating that Israel, since the start of the vaccination, is experiencing the rising phase of a ℛ_*c*_(*t*) fluctuating cycle, which is justified since the population is relaxing too soon also and taking the advantage of the benefit of an increasingly immunized community.

It has a best fit of (when excluding data since March 2, 2021 because ℛ_*c*_(*t*) has finally peaked):

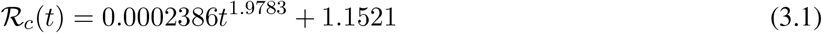

**Figure 3.2:**
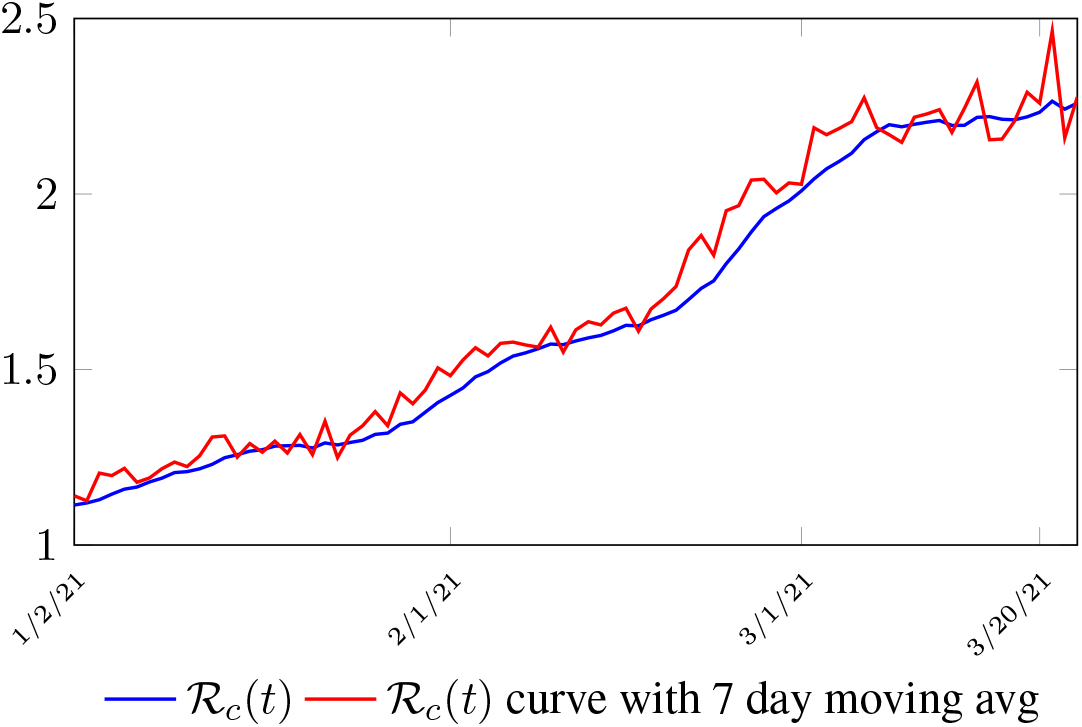
Israel’s ℛ_*c*_(*t*) curve

It is worth noting that the fit is an approximation of the final control reproduction number ℛ_*c*_(*t*) observed. Predictions can certainly deviates from reality, but it is forgivable as long as it is within the range of error of tolerance. The value of the intercept in equation (3.1) is the initial control reproduction number ℛ_*c*_ for Israel at the beginning of vaccination program. In fact, if the same public health measures were continually maintained and respected, Our result shows that Israel could have seen the last COVID-19 case on February 12, 2021 (Figure 3.3).As we have assumed in our previous work with fixed ℛ_*c*_(*t*). [33] Although under such scenario, the border should remain closed since herd immunity is not yet achieved despite zero indigenous infections. Based on Israel’s data, we know that COVID-19 variant strain has an intrinsic daily ℛ_0_ value of at least > 2.2, since the COVID-19 control ℛ_*c*_ value has arisen to 2.2 on the latest day of observation.

**Figure 3.3:**
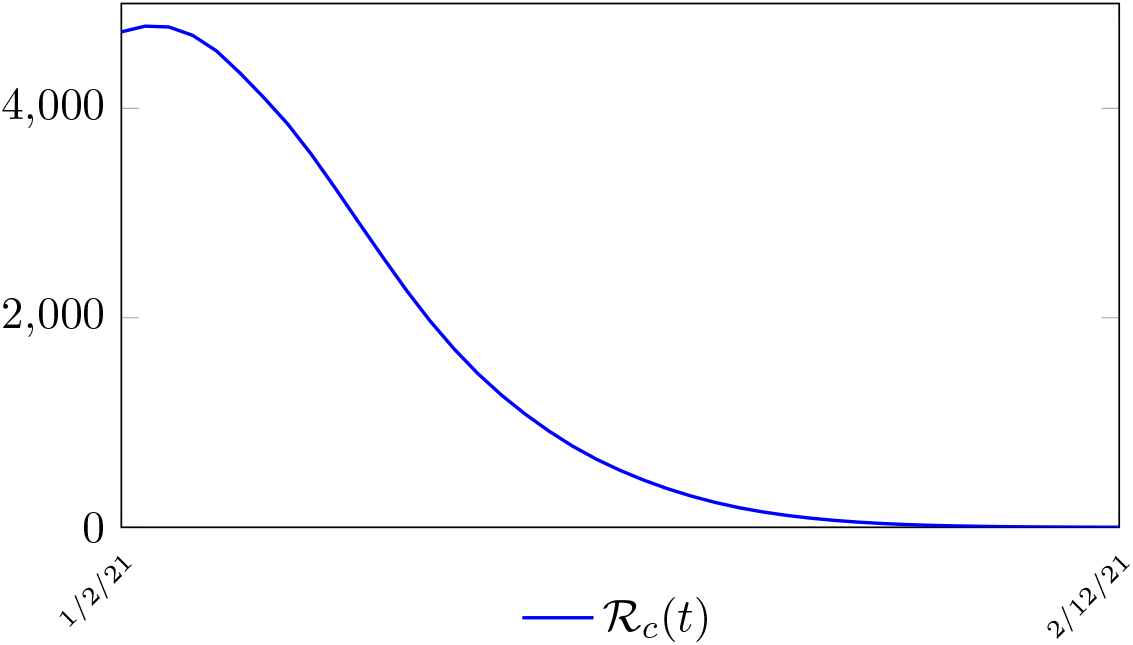
Israel’s predicted daily infection if control ℛ_*c*_ = 1.1521 remained.

Based on Israel’s data, *we cautiously conclude that COVID-19 variant strain has an intrinsic daily* ℛ_0_ *value of 2*.*2*, since the COVID-19 control ℛ_*c*_ value has arisen to 2.2 since March 2, 2020 and no longer increase despite a continued relaxation of measures and starts to fall short from our ℛ_*c*_(*t*) projection in magnitudes as we never seen before (Figure 3.4).

**Figure 3.4:**
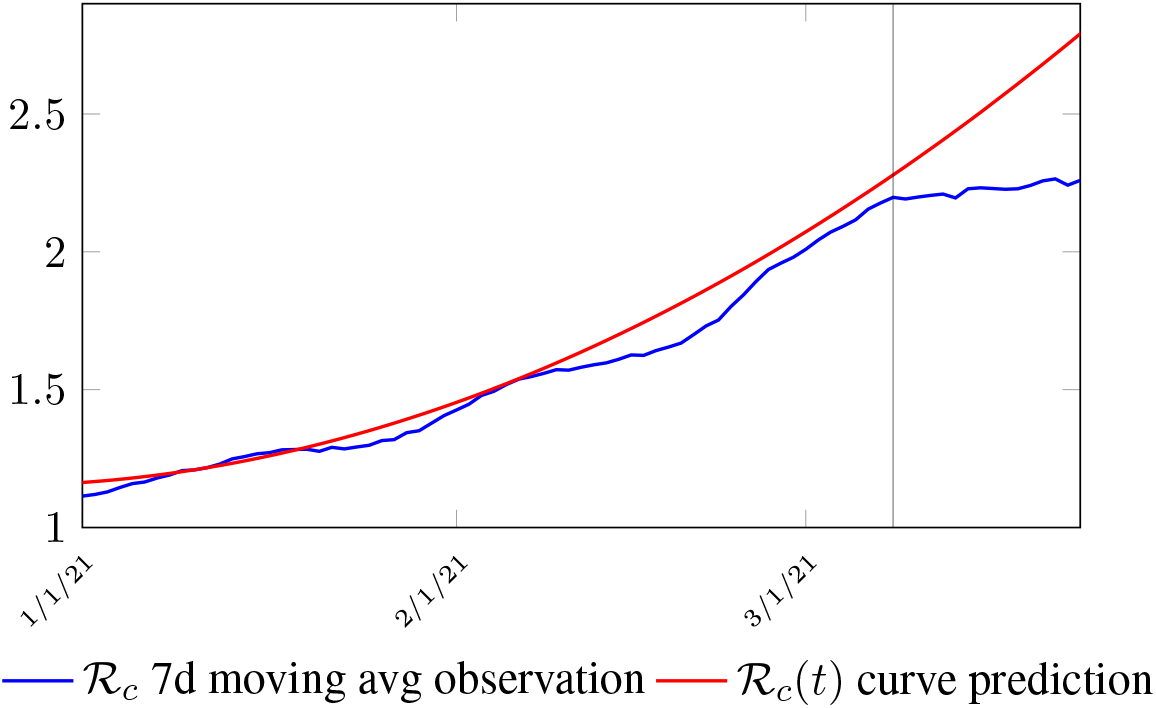
Observations starts to fall short from our ℛ_*c*_ (*t*) projection. Previous observations had also fall short from ℛ_*c*_ (*t*) projections, however, the gap has been much smaller, furthermore, the moving average was still increasing and not as a plateau as we observed now. The plateau starts at 3/8/21 indicated by the gray vertical divider.

In this case, we are lucky that the intrinsic ℛ_0_ value can be measured, in which the vaccination program lasts long enough so that there is enough time for ℛ_*c*_ (*t*) reaching its intrinsic ceiling value of ℛ _0_ (but it may not always be the case). In fact, if the prediction holds, Israel will report its last case by early April, so the end of time window for prediction is just 3 weeks away. In order to extrapolate future trend, we need compute the vaccination rate 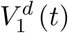 for 1 dose and vaccination rate 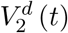 for 2 doses (Figure 3.5). Based on the past 1 month, no particular daily increase in single or double dose administration is observed [32]. Notice that by taking the 7 day moving average, one observe a 22 day delay between 1st dose and the second dose administration.

**Figure 3.5:**
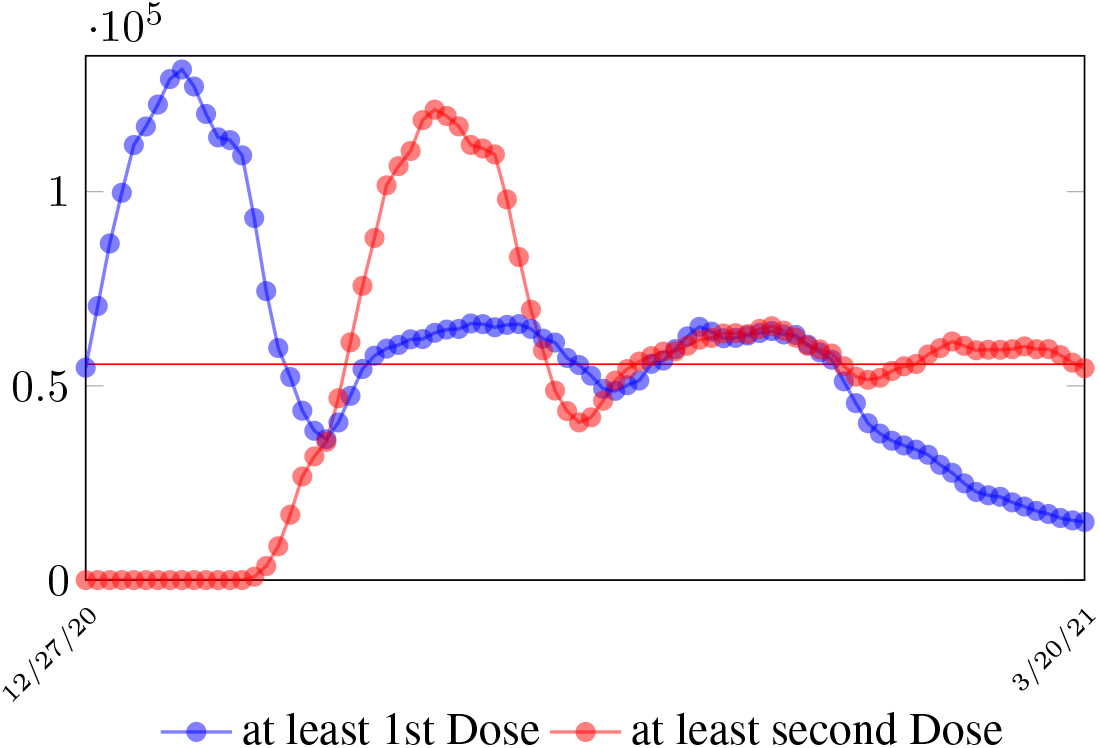
The observed Israel daily vaccination dosage administered for single and double doses during Feb 2021

As a result, we simply take the average dose administered during the month of February 2021 for single and double doses as the indicator for the vaccination rate of the future. So we have:

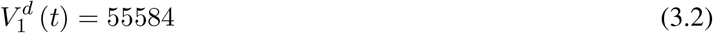

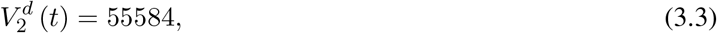

where the average dosage delivered for both 1st and second dose are identical except with a time delay. So we have Israel’s predictions in the upcoming months, the first simulation is assuming that the intrinsic ℛ_0_ ≥ℛ_*c*_ (*t*_*L*_). According to the simulation, when the last case is reported on April 29, of 2021, the final ℛ_*c*_ (*t*_*L*_) = 4.1. Based on our earlier discussion, if there exists *t*_0_ in which *t*_*F*_*≤ t*_0_ ≤*t*_*L*_ that the intrinsic ℛ_0_ is reached, then the predicted results for Israel would follow the other curves assuming ℛ_0_ = 2.5 and ℛ_0_ = 3 (Figure 3.6). The final ℛ_0_ shall only be determined based upcoming results, for which we have to patiently wait for now.

**Figure 3.6:**
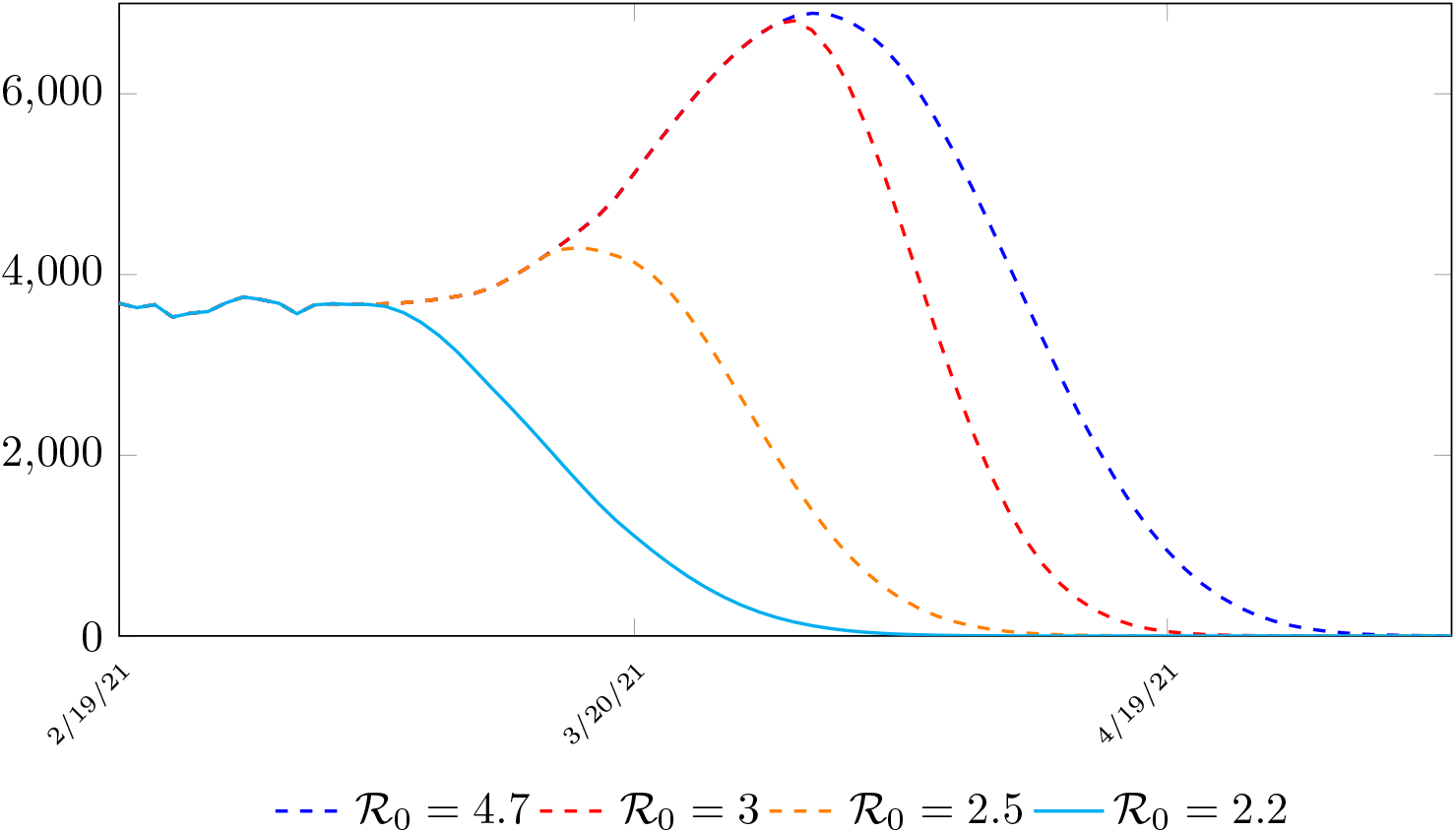
Israel predictions, left to right: ℛ_0_ = 2.2, ℛ_0_ = 2.5, ℛ_0_ = 3.0, and ℛ_0_ = 4.7. Its most likely that ℛ_0_ = 2.2. Other trajectories given higher possible ℛ_0_ is also shown. Notice that even for ℛ_0_ = 2.2 trajectory is somewhat higher than the actual number of infection observed. This is reasonable since the curve is based on 7 day moving average.

### 3.2 United States Projections

Figure 3.7 depicts the 7 day moving average on the daily vaccination for at least a single and double dosages administered in United States [34].

**Figure 3.7:**
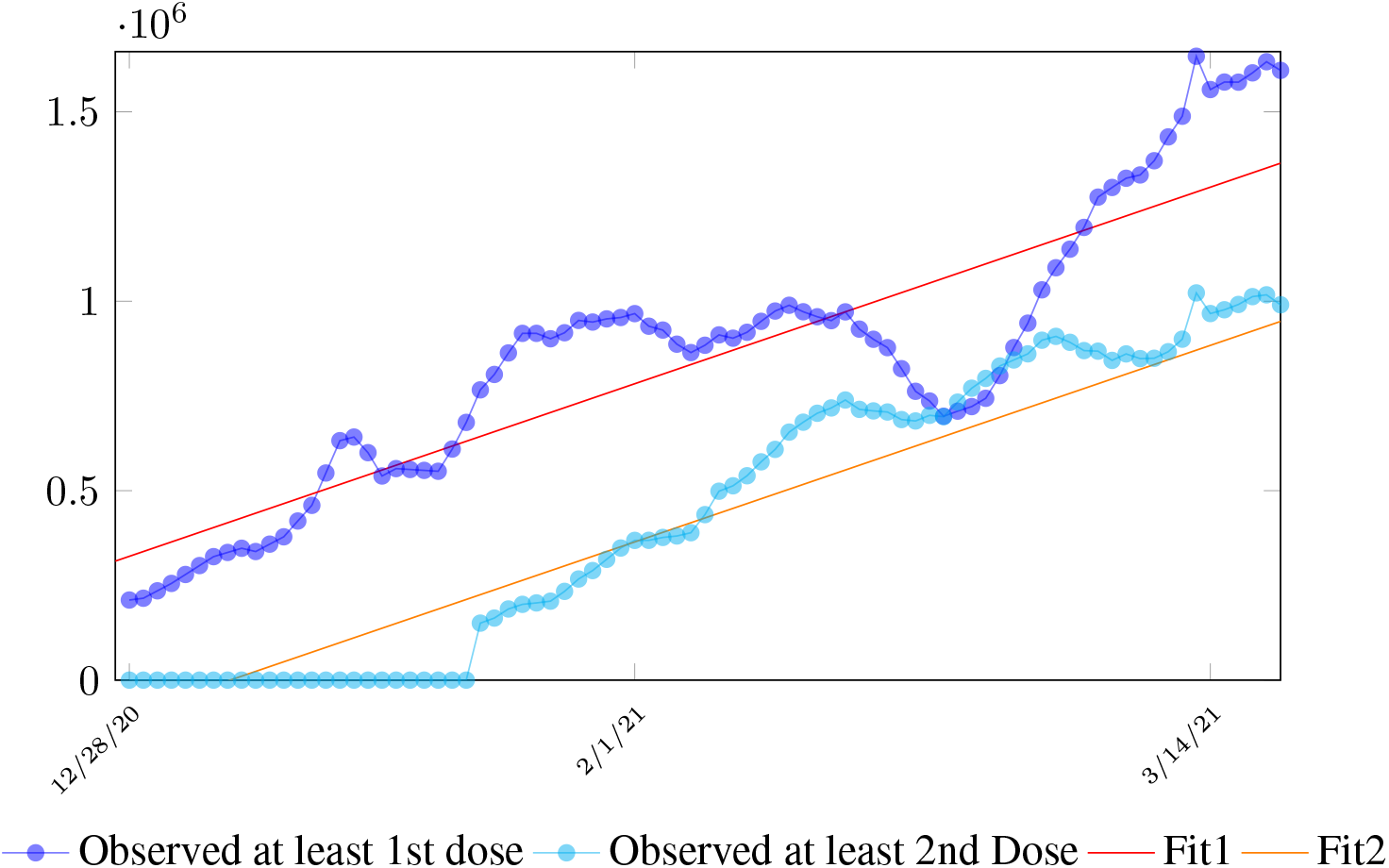
The 7 day moving average of US daily vaccination for dosage administered for at least 1 dose and its linear fit and at least 2 doses and its linear fit

We perform a linear fit for 1st dose given by

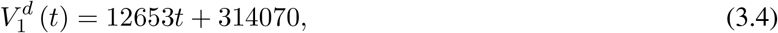

where the intercept 314070 is an over-estimation for vaccination administered record starting on Dec 28, 2020 and the second dose given by

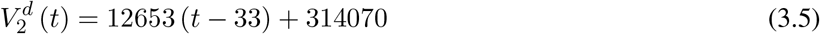

which is not the best linear fit with least *R*^2^, however, the idea behind such fit is simple. That is, there is a 1 month delay between all those had 1st dose and the second dose. Other linear fits with lower *R*^2^leads to higher slopes and eventually overtake the daily increase of those had at least 1 dose, which is logically impossible. We then compute *E* (*t*) and derived the immunization ratio of US as shown in Figure 3.8:

**Figure 3.8:**
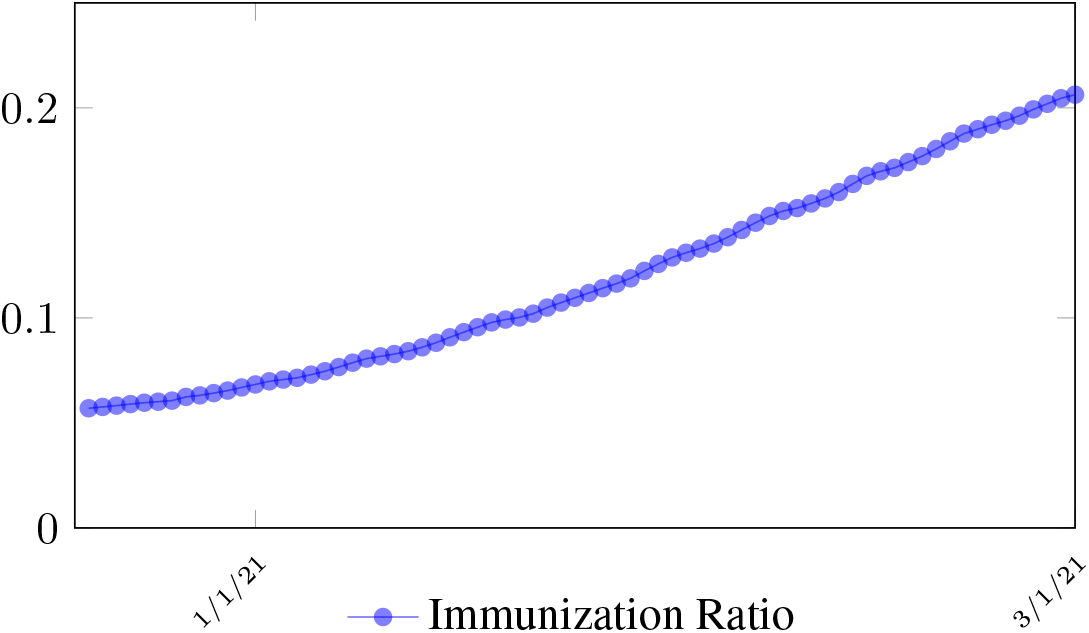
US effective immunization ratio

Figure 3.9 shows that the final ℛ_*c*_(*t*) curve is a rising one, indicating that US, since the start of the vaccination, is experiencing the rising phase of a ℛ_*c*_(*t*) fluctuating cycle, which is justified since the population is relaxing and taking the advantage of the benefit of an increasingly immunized community.

**Figure 3.9:**
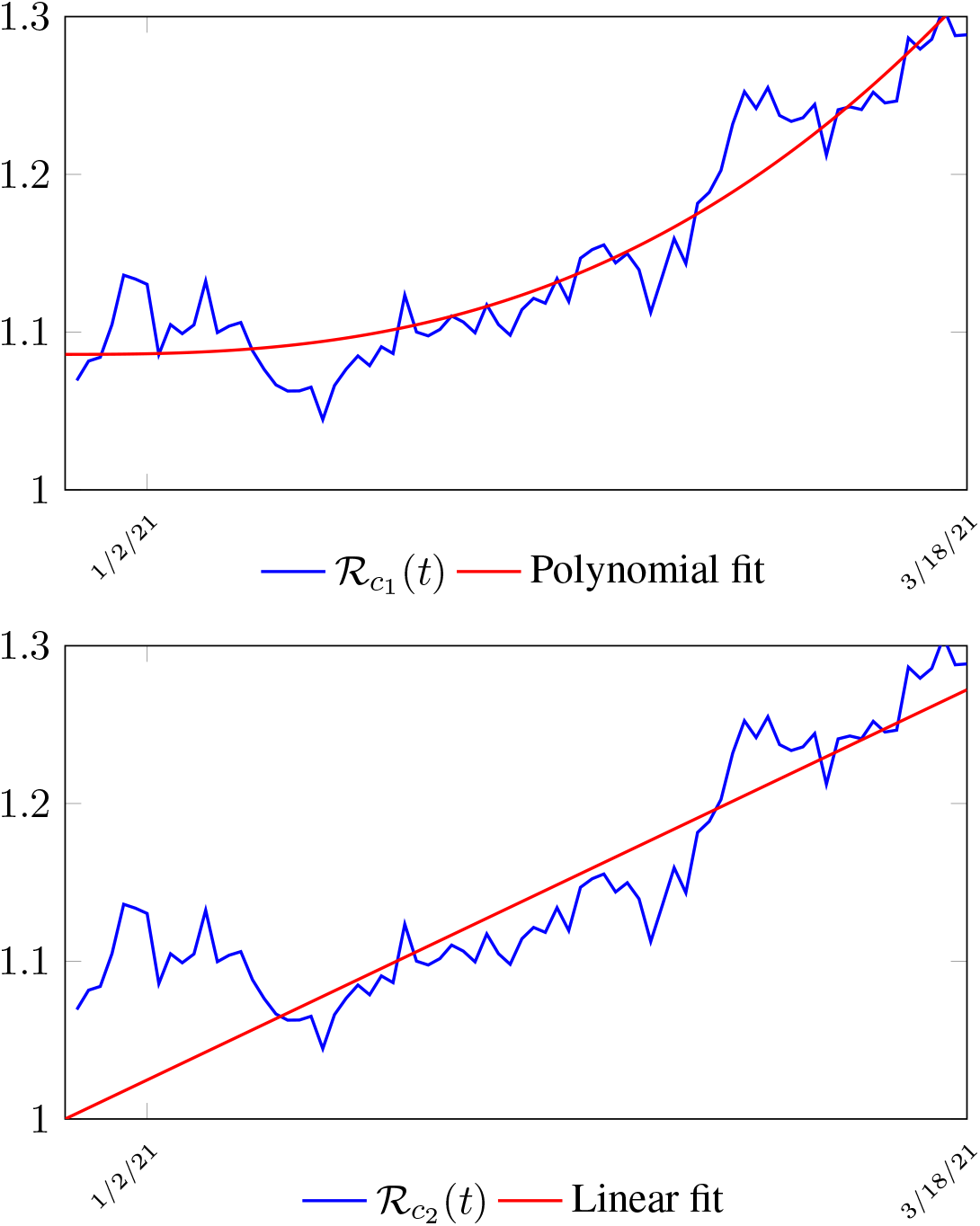
United States ℛ_*c*_(*t*) curve

We perform 2 fits for US, a polynomial and linear fit:

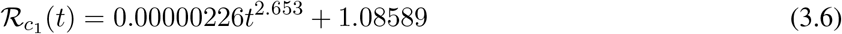

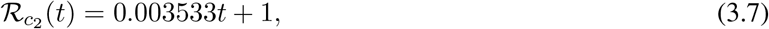

where the intercept 1.0828 and 1 is the assumed initial control ℛ_*c*_ value of US at the beginning of vaccination program. Along with ℛ_*c*_(*t*), we run the recursive simulation *S*(*t*). We find that depending on the ℛ_*c*_(*t*) curve, and assuming ℛ_0_ > 2.2, another surge of infection is imminent if the rising trend follows 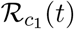. The vaccination rate will be too slow to catch up with the relaxation of human behaviors returning to normalcy, despite daily vaccination increase at a linear rate. With an initial infection size of 30,081,657 on March 14, 2021, additional infection of 6,101,698 and cumulative effective vaccination size of 243,095,373 yields a cumulative effective immunization ratio *E*(*t*) = 0.867 on June 30, 2021 when ℛ_*c*_(*t*) = 3.427. Assuming ℛ_0_ = 2.2, another surge still ensues but with somewhat faster decline, then additional infection of 5,624,152 and cumulative effective vaccination size of 216,944,672 yields a cumulative effective immunization ratio *E*(*t*) = 0.784 on Jun 19, 2021 when ℛ_*c*_(*t*) = 2.2. If the rising trend follows 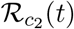, one should observe a steady decreases of cases with a linear gradual return to normalcy with no additional surge and rebound. In that case, the last case is reported on May 15, 2021 when ℛ_*c*_(*t*) = 1.495 with additional infection of 706,952 and cumulative effective vaccination size of 143,813,093 yields a cumulative effective immunization ratio *E*(*t*) = 0.542.

In all scenarios, zero additional cases are reported well before *t*_*L*_, the assumed last day of total vaccination delivery is made when *E* (*t*_*L*_) ≤1. If human collective behavior changes in response to zero infection case achieved, and assuming that

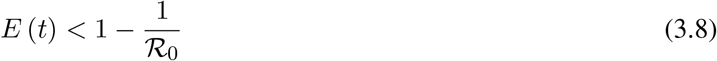

That is, the endemic equilibrium is still higher than the cumulative effective immunization ratio achieved, then it is inevitable resurgence ensues with the imports of overseas cases, and a sudden discontinuity of positive jump on the ℛ_*c*_(*t*) over a short period of time. As a result, vaccination delivery must be continued and restrictive measures respected even if zero cases are reported. Under 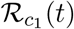 with a cumulative effective immunization ratio *E*(*t*) = 0.867 and assuming in the worst case scenario in which ℛ_0_ → ∞, then another 16 days of vaccination is required:

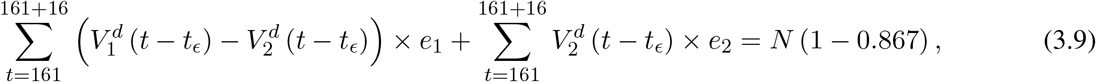

where *t* = 161 is the number of days passed since Dec 27, 2020 when zero case is reported. That is, the pandemic ends in the US by July 16, 2021 if citizens collectively continue to respect measures in place and very gradually restores to normalcy even well after the last case is reported in June 30, 2021. Under 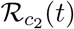 scenario, which results a cumulative effective immunization ratio *E*(*t*) = 0.542 when the last case is reported in May 15, then another 64 days of vaccination is required:

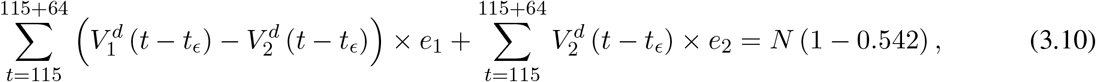

where *t* = 115 is the number of days passed since Dec 27, 2020 when zero case is reported. That is, the pandemic officially ends in the US by July 18, 2021 when 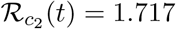 if citizens collectively continue to respect measures in place and very gradually restores to normalcy even well after the last case is reported in May 15, 2021. Only when one assumes that 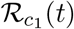 with a cumulative effective immunization ratio *E*(*t*) = 0.784 and assuming ℛ_0_ = 2.2, and 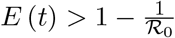, then normalcy can be immediately restored as:

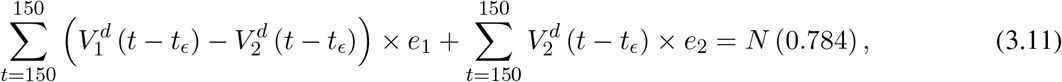

where *t* = 150 is the number of days passed since Dec 27, 2020 when zero case is reported. That is, the pandemic ends in the US by June 19, 2021. Although, in this case, sporadic new imported cases can still be reported and causes local spread and quickly terminates without creating resurgence. The analysis shows that the battle against COVID-19 in US is no easier than 2020, a chance of resurgence is not only possible but possibly imminent unless people collectively remain cautious throughout the year despite a continually implemented vaccination program on a daily bases. That is, the administration of vaccine and even zero reported cases is by no means of a sign of relaxation and anxiety of returning to a complete normalcy, as long as the effective immunization ratio *E* (*t*) *<* 1 and ℛ_0_ value remains inconclusive.

### 3.3 United Kingdom Projections

During the month of March 2021, no particular daily increase in single dose administration is observed in UK (Figure 3.11) [35]. For the second dose, there is an increasing trend, but we shall conservatively assume that the trend will simply flat out. More interestingly, UK second dose does not follow the trend typically observed in other nations where the second dose administration typically follows the 1st dose with a 2 to 3 weeks delay.

**Figure 3.10:**
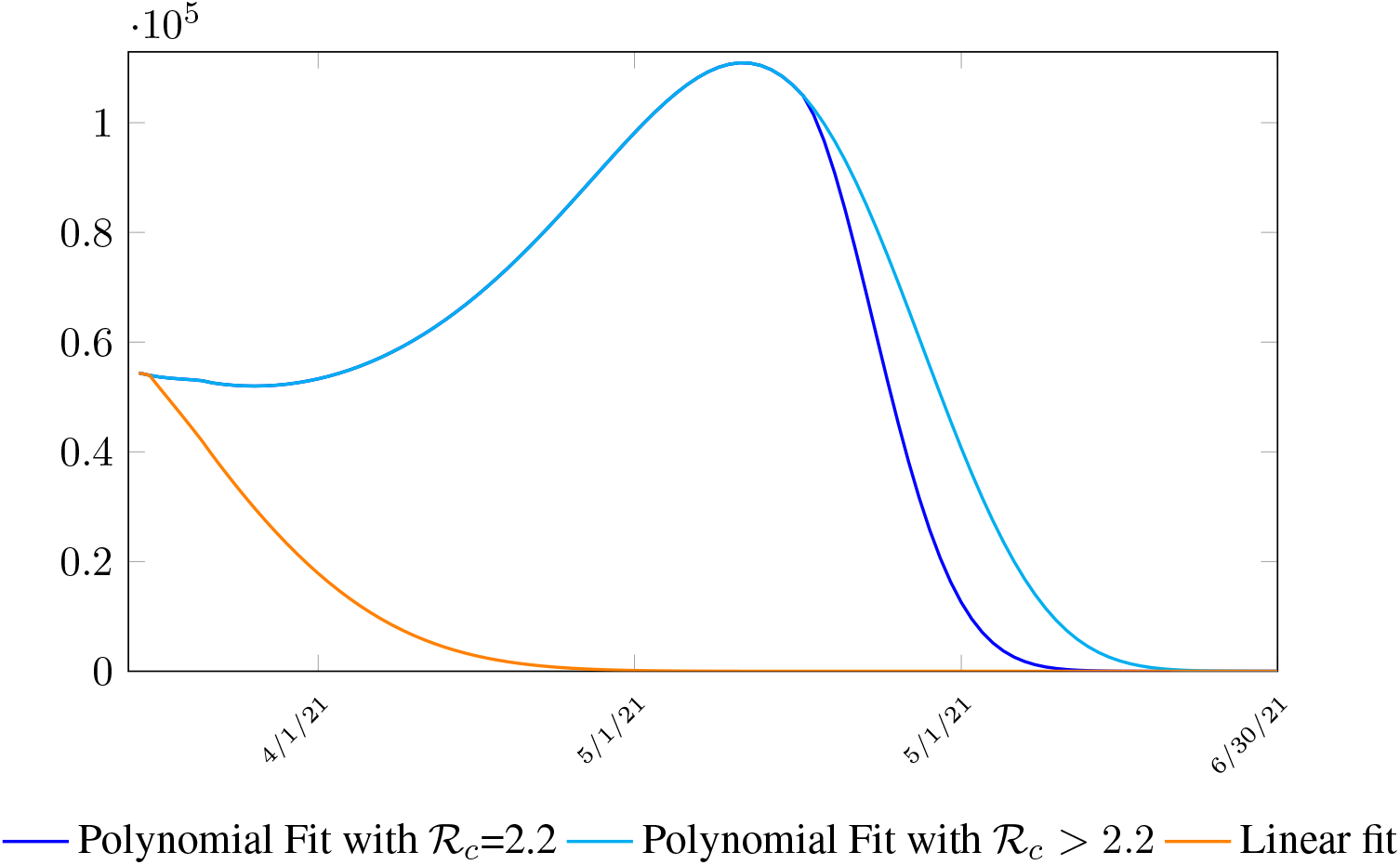
US observed number of cases under 3 different trajectories

**Figure 3.11:**
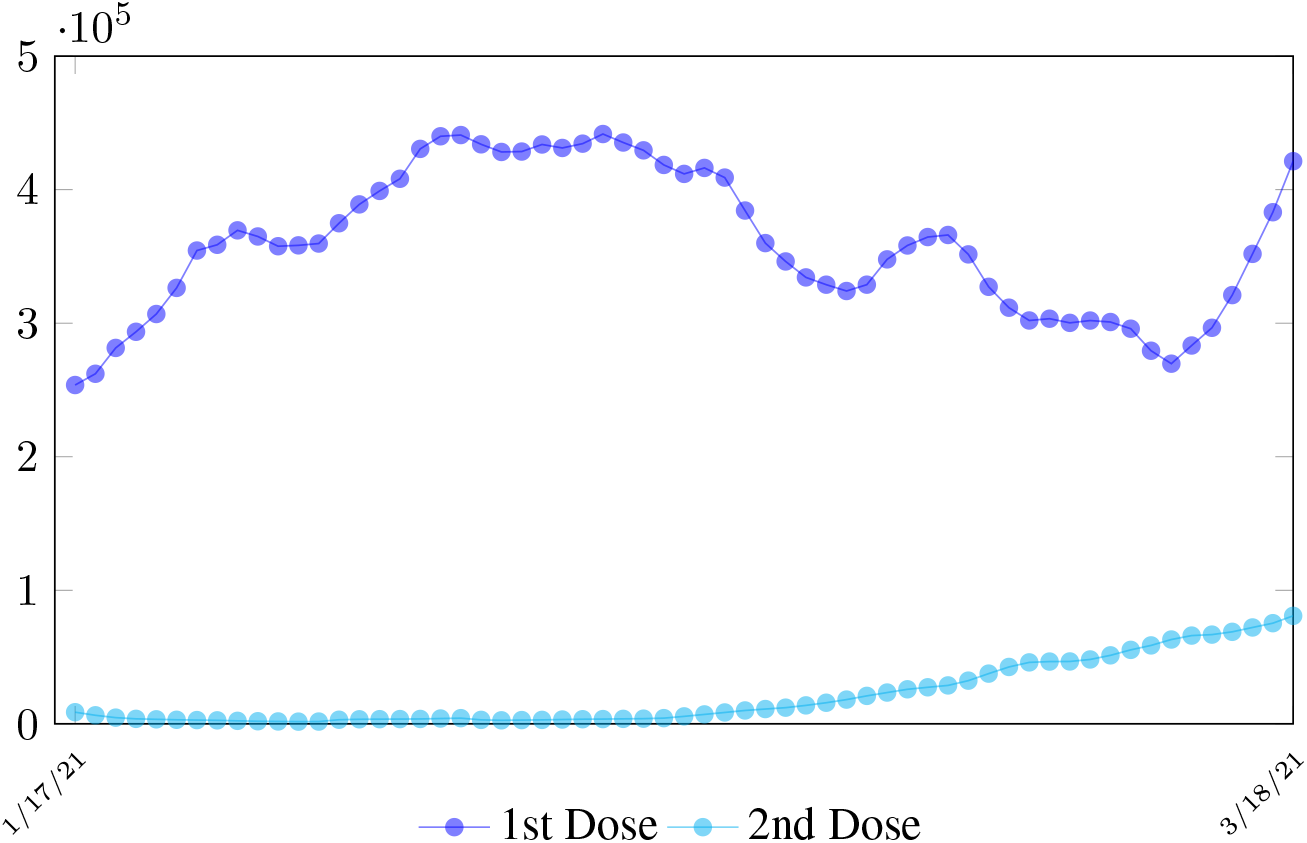
The observed UK daily vaccination dosage administered for at least single and at least double doses during late January to early March 2021

As a result, we simply take the average daily dose administered during late January to early March 2021 for single dose and average daily dose administered during the last 2 weeks of late February and early March 2021 for double doses as the indicator for the vaccination rate of the future. So we have:

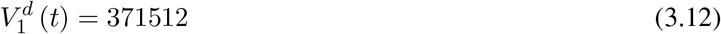

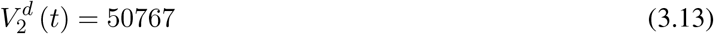

We then compute *E* (*t*) and derived the immunization ratio of UK as given in Figure 3.12

**Figure 3.12:**
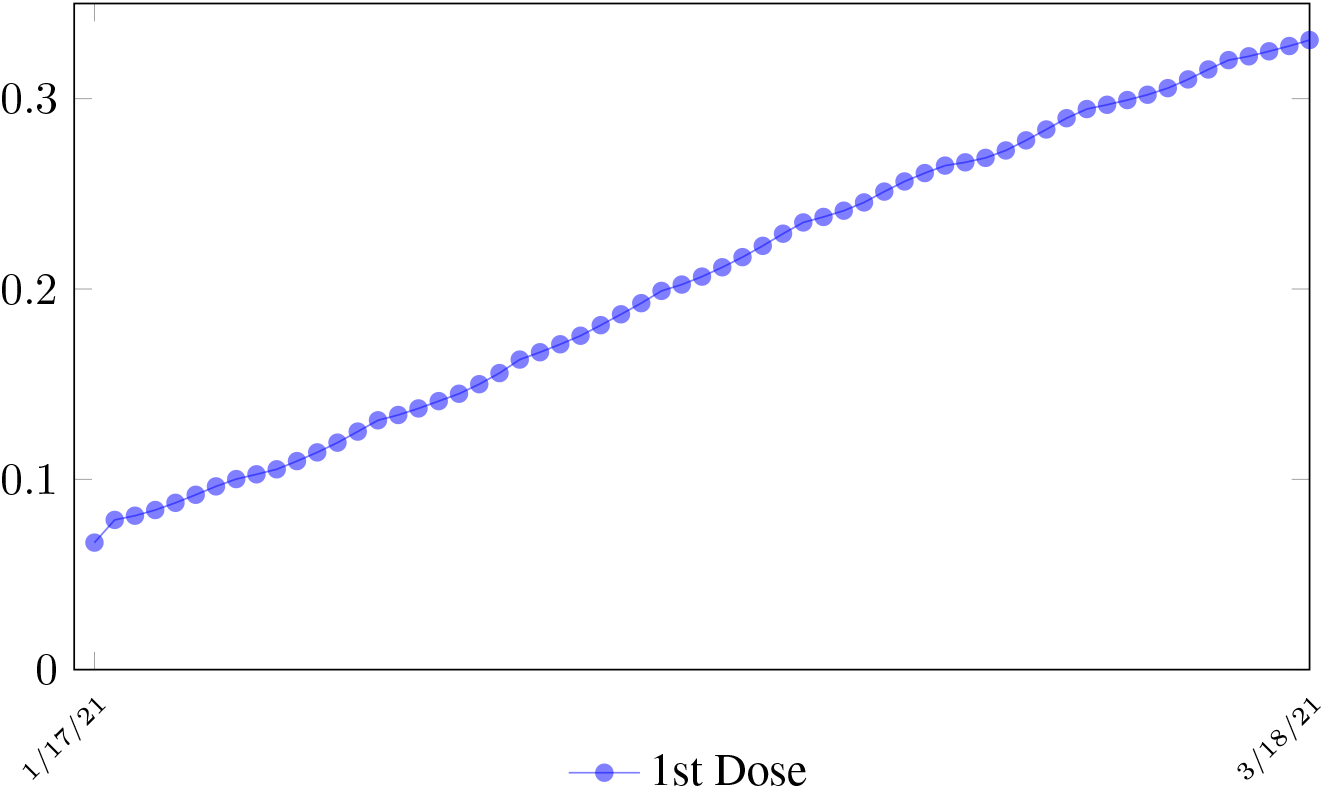
UK effective immunization ratio

We perform 2 fits for UK, a polynomial and linear fit:

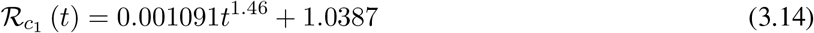

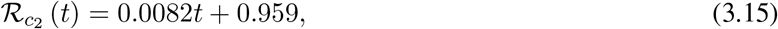

where the intercept 1.03 is the initial control ℛ_*c*_ value of UK at the beginning of vaccination program (Figure 3.13). Along with ℛ_*c*_ (*t*), we run the recursive simulation *S* (*t*). We find that depending on the ℛ_*c*_ (*t*) curve, and assuming ℛ_0_ > 2.2, no additional surge of infection is observed if the rising trend follows 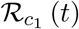. The vaccination rate is faster than the relaxation rate of human behaviors returning to normalcy. With an initial infection size of 4,285,684 on 3/19/21, [36, 37] additional infection of 183,799 and cumulative effective vaccination size of 38,115,429 yields a cumulative effective immunization ratio *E* (*t*) = 0.735 on June 17, 2021 (Figure 3.14) when ℛ_*c*_(*t*) = 2.766. If the rising trend follows 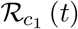, one should also observe a faster steady decreases of cases with a linear gradual return to normalcy with no additional surge and rebound. In that case, the last case is reported on May 25, 2021 when ℛ_*c*_(*t*) = 2.017 yields a cumulative effective immunization ratio *E* (*t*) = 0.6334.

**Figure 3.13:**
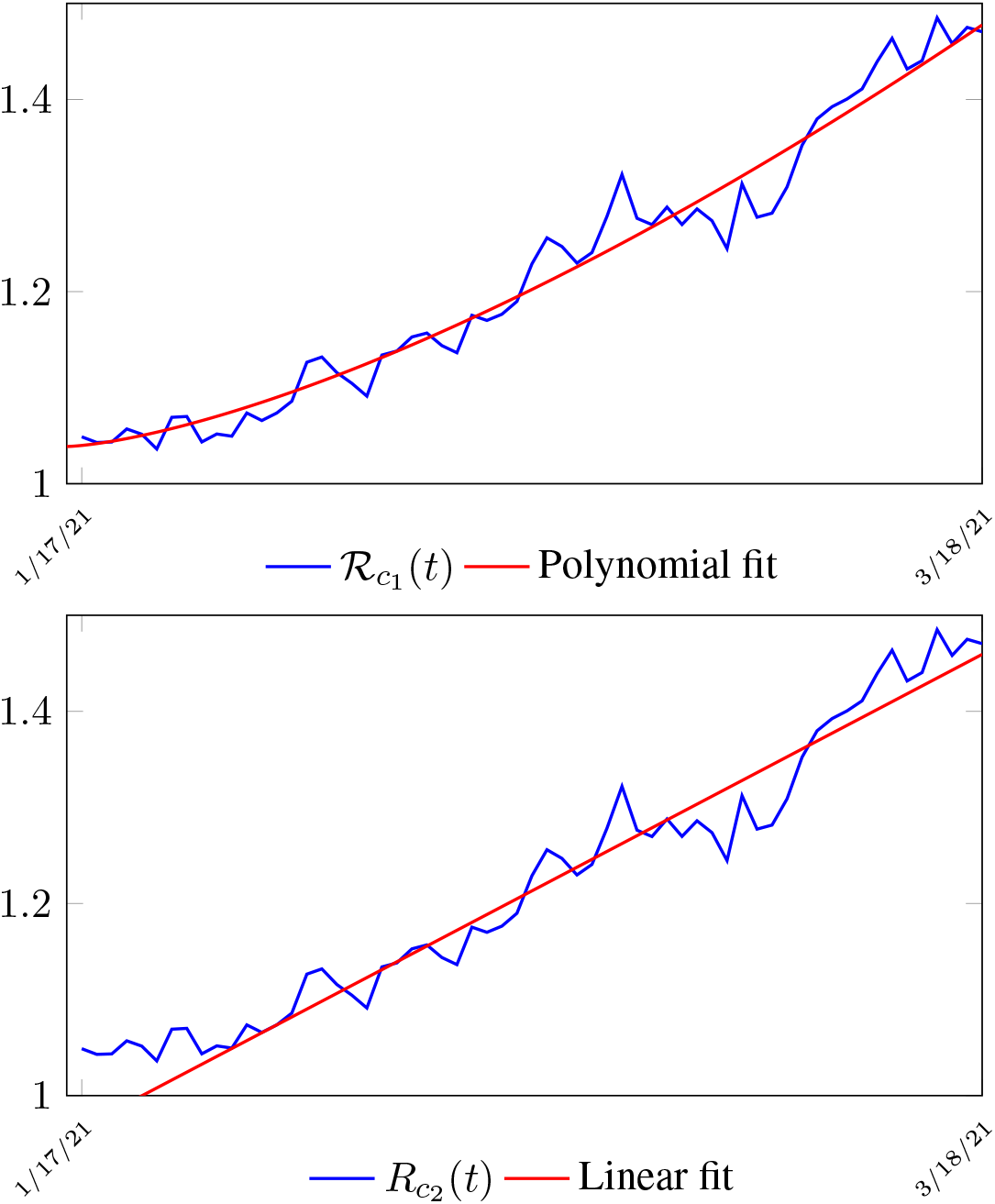
United Kingdom *R*_*c*_(*t*) curve

**Figure 3.14:**
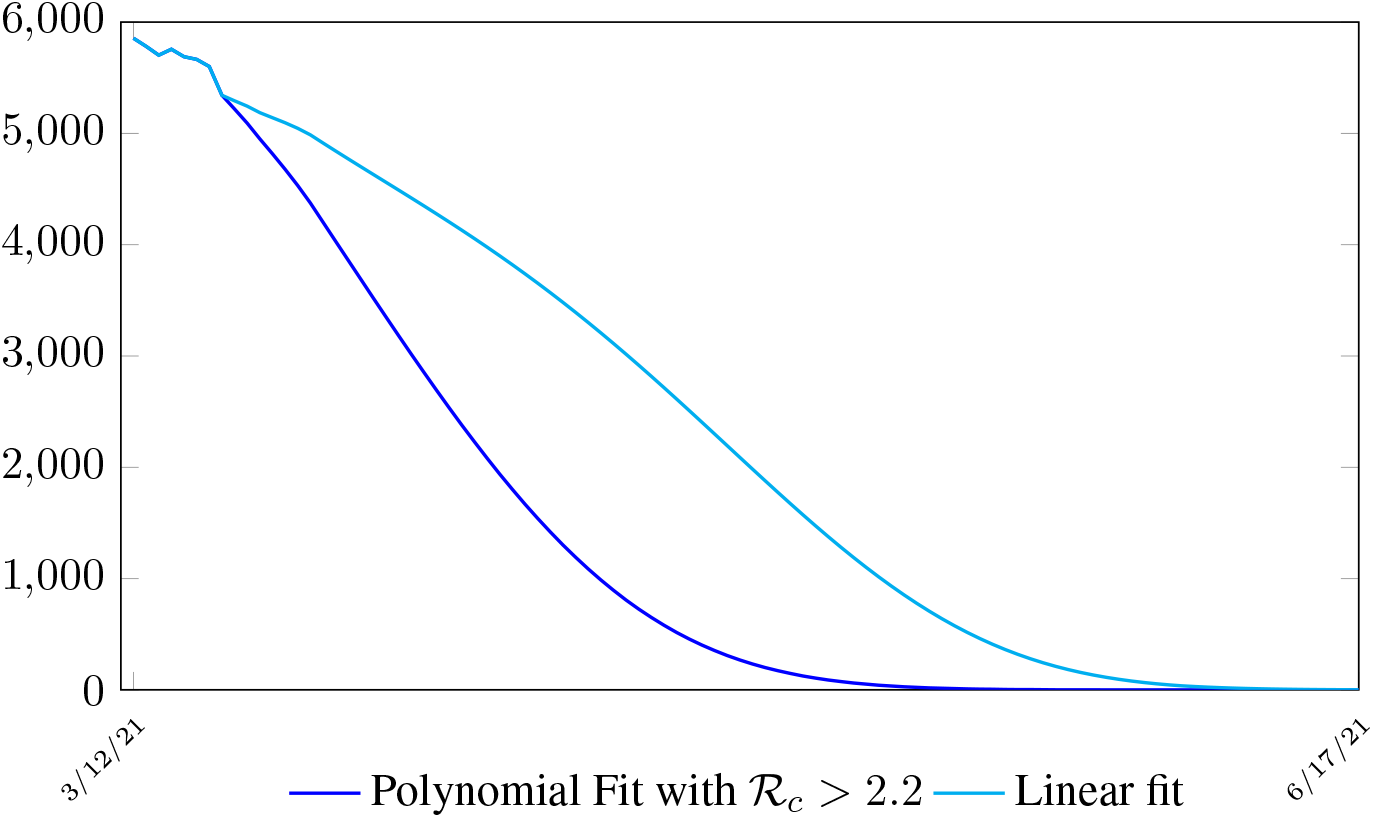
UK observed number of cases under 2 different ℛ_*c*_ (*t*) curv

In all scenarios, zero additional cases are reported well before *t*_*L*_, the assumed last day of total vaccination delivery is made when *E* (*t*_*L*_) ≤ 1. If human collective behavior changes in response to zero infection case achieved, and assuming that

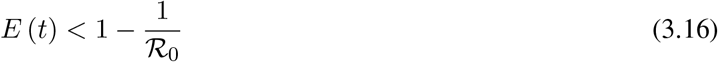

That is, the endemic equilibrium is still higher than the cumulative effective immunization ratio achieved, then it is inevitable resurgence ensues with the imports of overseas cases, and a sudden discontinuity of positive jump on the ℛ_*c*_ (*t*) over a short period of time. As a result, vaccination delivery must be continued and restrictive measures respected even if zero cases are reported. Under 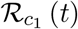 with a cumulative effective immunization ratio *E* (*t*) = 0.735 and assuming in the worst case scenario in which ℛ_0_ → ∞, then another 16 days of vaccination is required:

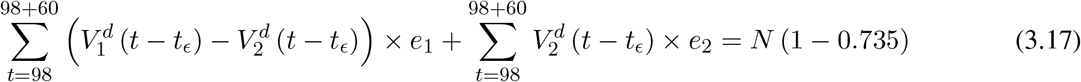

Whereas *t* = 98 is the number of days passed since Mar 12, 2021 when zero case is reported. That is, the pandemic ends in the UK no later than September 24, 2021 if citizens collectively continue to respect measures in place and very gradually restores to normalcy even well after the last case is reported in June 17, 2021.

Under 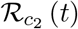 scenario, which results a cumulative effective immunization ratio *E* (*t*) = 0.6334 when the last case is reported in May 15, then another 64 days of vaccination is required:

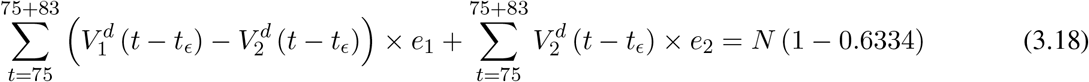

Whereas *t* = 75 is the number of days passed since Mar 12, 2021 when 0 case is reported. That is, the pandemic officially ends in the UK by 9/24/21 when 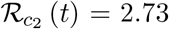 if citizens collectively continue to respect measures in place and very gradually restores to normalcy even well after the last case is reported in 5/25/21.

The reason that in both cases the pandemic ends on the same date is that the difference between the infections under 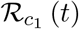 and 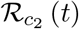 with the latest observed date until the 0 reported cases into the future based on simulation is minimal so that

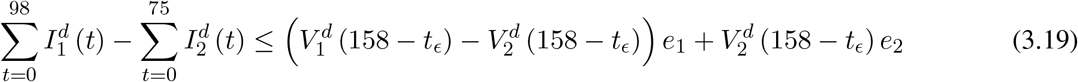

it is less than the daily vaccination rate at day 158 −*t*_*ϵ*_, so any difference is compensated within a single day. In general, a pandemic ends *n*_2_ days faster under 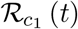 when *t*_*E*1_ is the last day of infection reported under 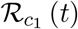. *t*_*E*1_> *t*_*E*2_, *n*_1_ is the additional days required for *E* (*t*) = 1 under 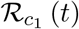 after *t*_*E*1_. *n*_1_+ *n*_2_ is the additional days needed for *E* (*t*) = 1 under 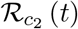 after *t*_*ϵ*2_.

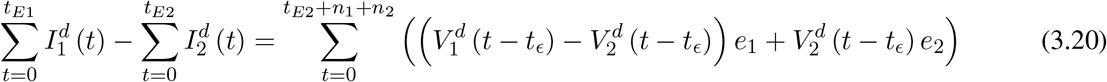

### 3.4 Serbia Projections

Based on all historical records, no particular daily increase in single or double dose administration is observed in Serbia, [38] and the second dose administration lags behind the 1st by 17 days, as typically observed in Israel and US as in Figure 3.15.

**Figure 3.15:**
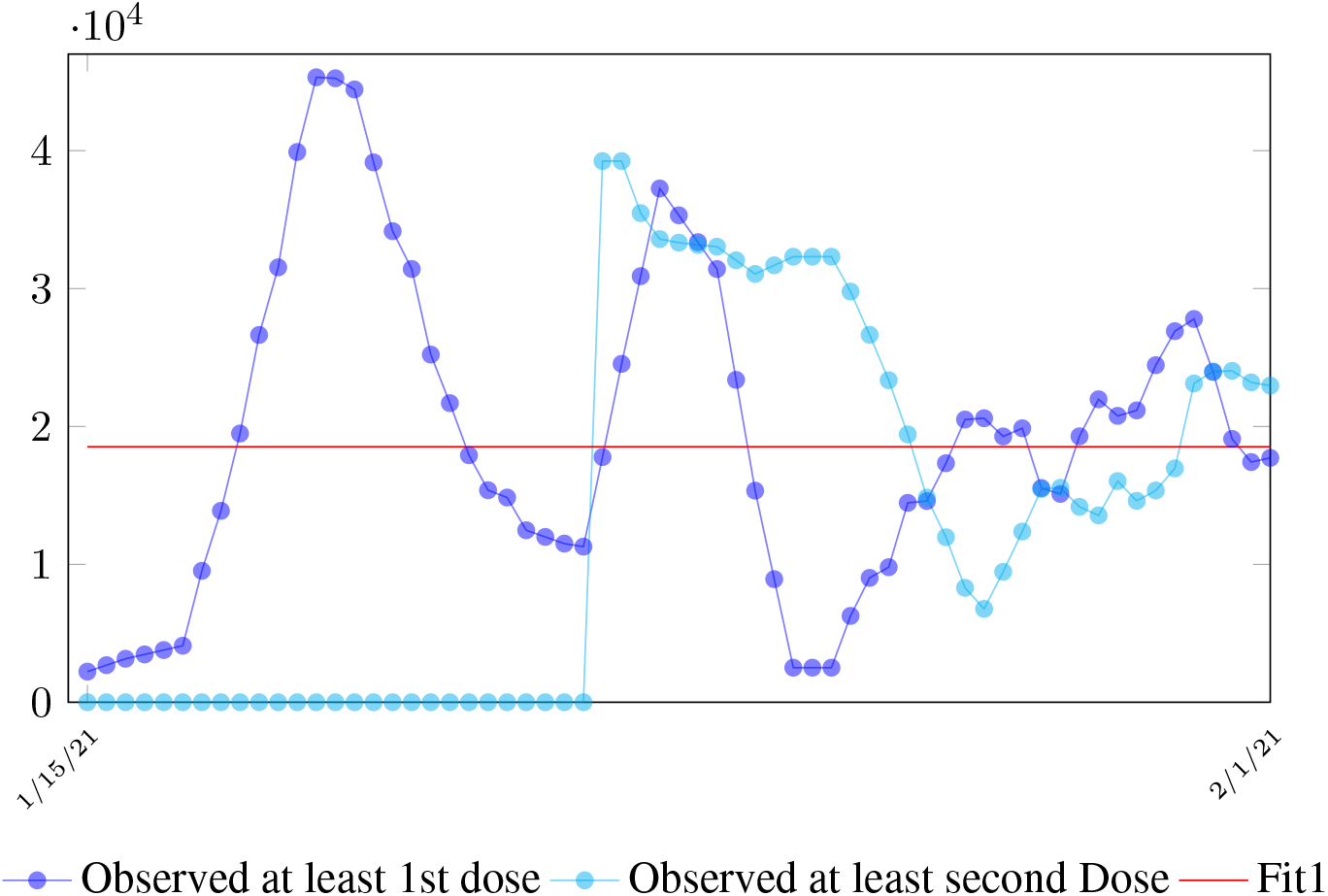
The 7 day moving average of Serbia daily vaccination for dosage administered for at least 1 dose and at least 2 doses

As a result, we simply take the average daily dose administered during the entire 2021 for single dose and average daily dose administered during the last 1 month from Feb to March 2021 for double doses as the indicator for the vaccination rate of the future. So we have:

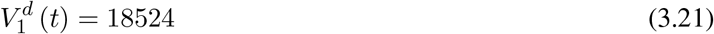

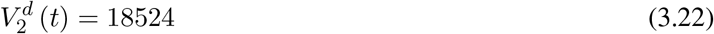

We then compute *E* (*t*) and derived the immunization ratio of Serbia as given in (Figure 3.16)

**Figure 3.16:**
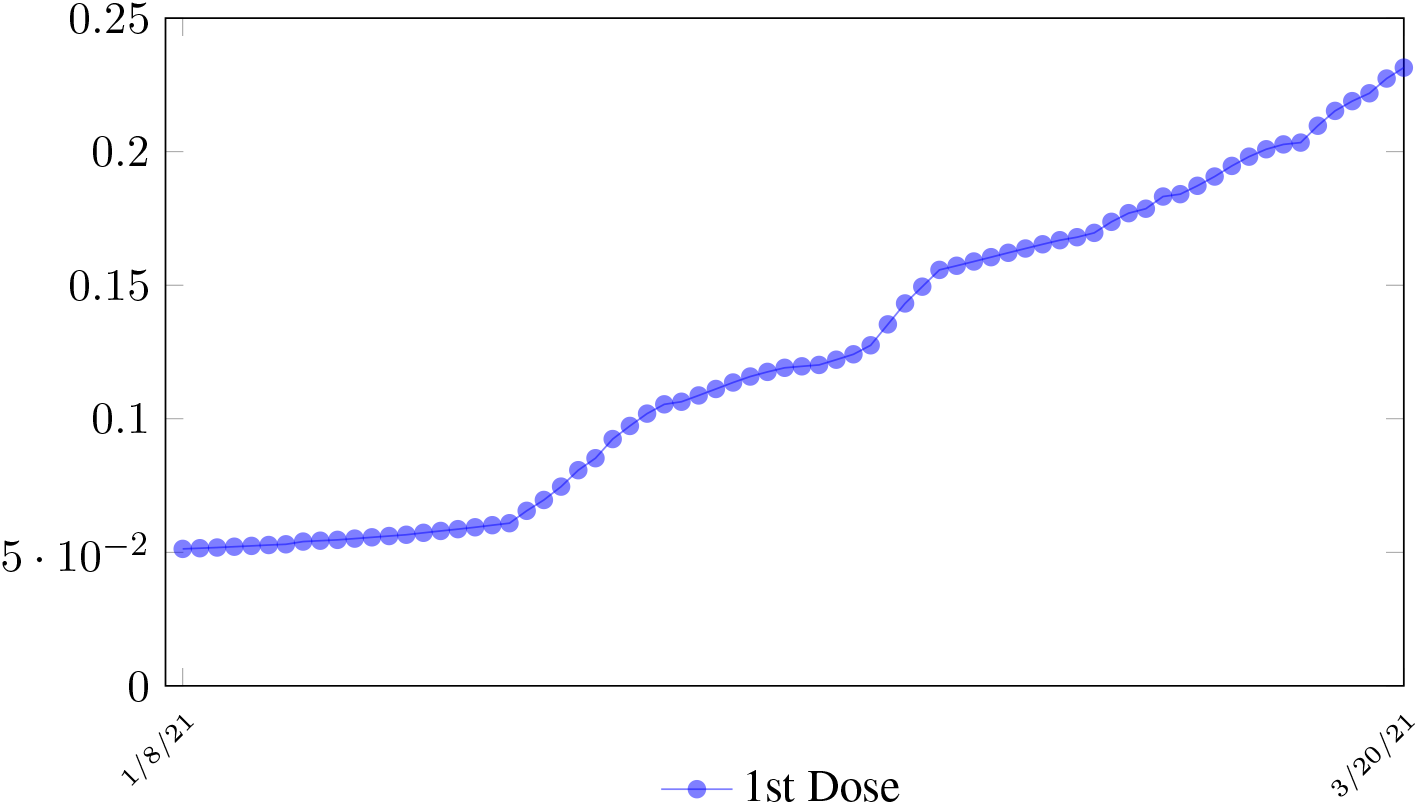
Serbia effective immunization ratio

We perform 2 fits for Serbia, a polynomial and linear fit, but polynomial fit is actually sub-linear so we have a single linear fit:

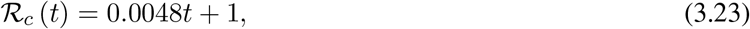

where the intercept 1 is the initial control ℛ_*c*_ value of Serbia at the beginning of vaccination program.

Along with ℛ_*c*_ (*t*), we run the recursive simulation *S* (*t*). We find that depending on the ℛ_*c*_ (*t*) curve, and assuming ℛ_0_ > 2.2, a small surge of infection is observed if the rising trend follows ℛ_*c*_ (*t*) (Figure 3.17). The vaccination rate is somewhat slower than the relaxation rate of human behaviors returning to normalcy. With an initial infection size of 546,896 on 3/20/21, [39] [40] additional infections of 357,628 and cumulative effective vaccination size of 6,561,999 yields a cumulative effective immunization ratio *E* (*t*) = 0.5817 on July 18, 2021 when ℛ_*c*_(*t*) = 1.9. (Figure 3.18).

**Figure 3.17:**
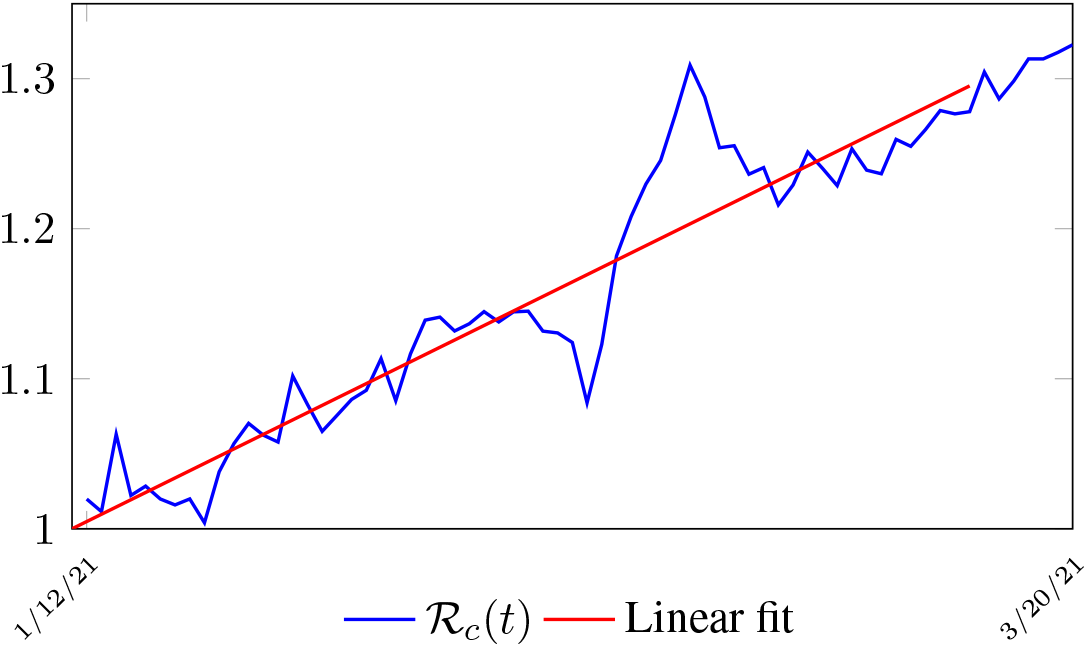
Serbia ℛ_*c*_ (*t*) curve

**Figure 3.18:**
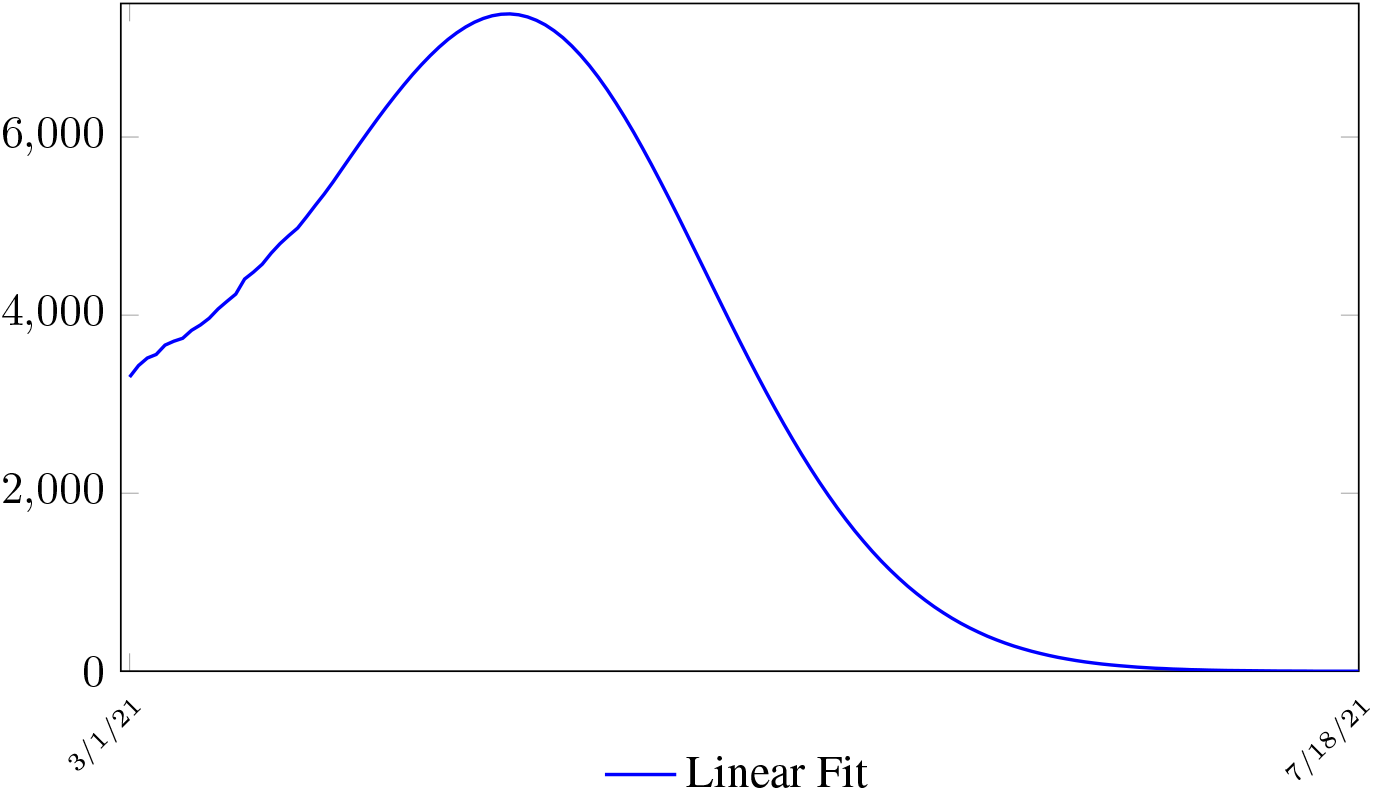
Serbia observed number of cases under ℛ_*c*_(*t*) curve

zero additional cases are reported well before *t*_*L*_, the assumed last day of total vaccination delivery is made when *E*(*t*_*L*_) ≤ 1. If human collective behavior changes in response to 0 infection case achieved, and assuming that

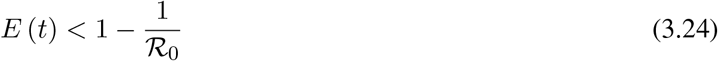

That is, the endemic equilibrium is still higher than the cumulative effective immunization ratio achieved, then it is inevitable resurgence ensues with the imports of overseas cases, and a sudden discontinuity of positive jump on the ℛ_*c*_ (*t*) over a short period of time. As a result, vaccination delivery must be continued and restrictive measures respected even if zero cases are reported. Under ℛ_*c*_ (*t*) with a cumulative effective immunization ratio *E* (*t*) = 0.5871 and assuming in the worst case scenario in whichℛ _0_→ ∞, then another 164 days of vaccination is required:

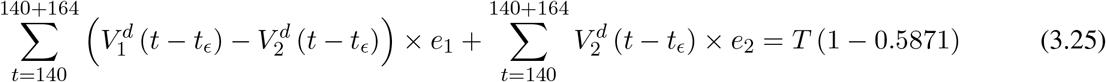

Whereas *t* = 140 is the number of days passed since Mar 1st, 2021 when zero case is reported. That is, the pandemic ends in Serbia no later than 12/29/21 if citizens collectively continue to respect measures in place and very gradually restores to normalcy even well after the last case is reported in 7/18/21.

## 4 Discussion

The relationship between *t*_*ϵ*_, the delay of the onset of antibodies after immunization, and *e*_1_, the efficiency for inducing antibodies based immunity with at least one dose requires further analysis. In our paper, we assumed there is a 8 days delay for the onset of antibodies after immunization. However, other literature suggests a range of delays [29]. There are two possible assumptions. First, *e*_1_ can remain fixed while the delay is lengthened to *t*_*ϵ*_ + *t*_*d*_. In this case, the final immunity ratio *E* (*t*) decreases because *V*_1_(*t* − (*t*_*ϵ*_ + *t*_*d*_)):

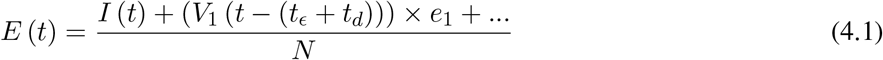

Second, *e*_1_ is increased to *e*_1_+ *e*_*d*_ while the delay is lengthened to *t*_*ϵ*_ + *t*_*d*_. In this case, it suggests that our previous delay was too short to cover all possible cases with induced immunity. The final *E* (*t*) is then depending on the magnitude of *e*_*d*_ and *t*_*d*_ as:

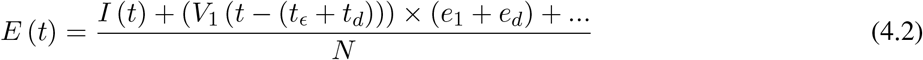

The second case led to us to the conclusion that both models are approximations. In reality, the number of immunized increases in a time dependent manner. In the simplest form, the cumulative number of people with induced immunity grows linearly with each passing day from a pool of people initially vaccinated. If the daily newly induced is constant, then a uniform distribution spread across time can model daily changes. Then, weighted average delay *t*_*ϵ*_ is:

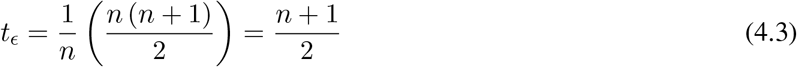

assuming in *n* days all possible induced immunization is realized and each day shares 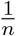 out of all induced. One can easily see in that case the final *t*_*ϵ*_ should be the median of *n* days registered. In reality, each passing day likely to have weighted proportion, and non-uniform distribution such as normal distribution spread across time can probably model daily changes more accurately. In such case, the cumulative number of people with induced immunity grows according to a logistic function. In general, assume there is a function *F* (*V*_1_(*t*), *t* + *t*_*i*_) that takes initial size of people vaccinated on day *t* and given a subsequent day *t* + *t*_*i*_, returns the number of people with induced immunity on day *t* + *t*_*i*_. Then the weighted average delay *t*_*ϵ*_ can be computed as:

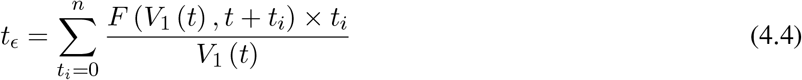

However, weighted average delay *t*_*ϵ*_ is yet another approximation attempt at its best. To truly represent the number of people with induced immunity one has to construct a (*t*_*L*_ − *t*_*F*_) (*t*_*L*_ − *t*_*F*_ + *n* + 1) sized matrix *A* as:

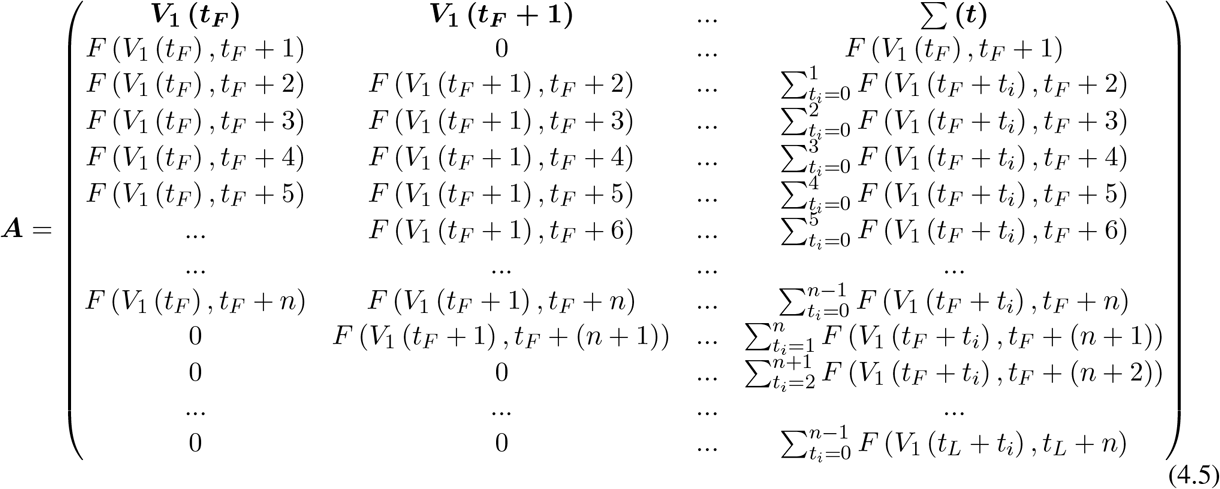

Whereas the final results are tallied under the last row ***A*** [*t*_*L*_ *—t*_*F*_ + 1], and *V*_1_ (*t*) × *e*_1_ can be re-defined as the cumulative of all with induced immunity from vaccinations from all days:

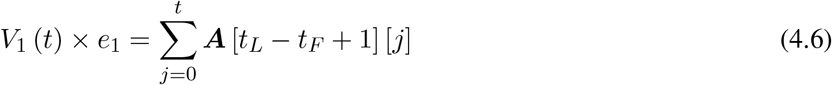

and daily effective vaccination can be defined as:

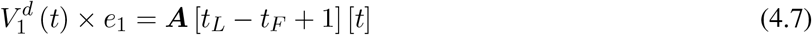

Correspondingly, similar matrix can be constructed for the second dose. This certainly leaves room for future work to make predictions more precise.

Finally, the relationship diagram for deriving ℛ_*c*_, the control ℛ_0_ can be illustrated as below:

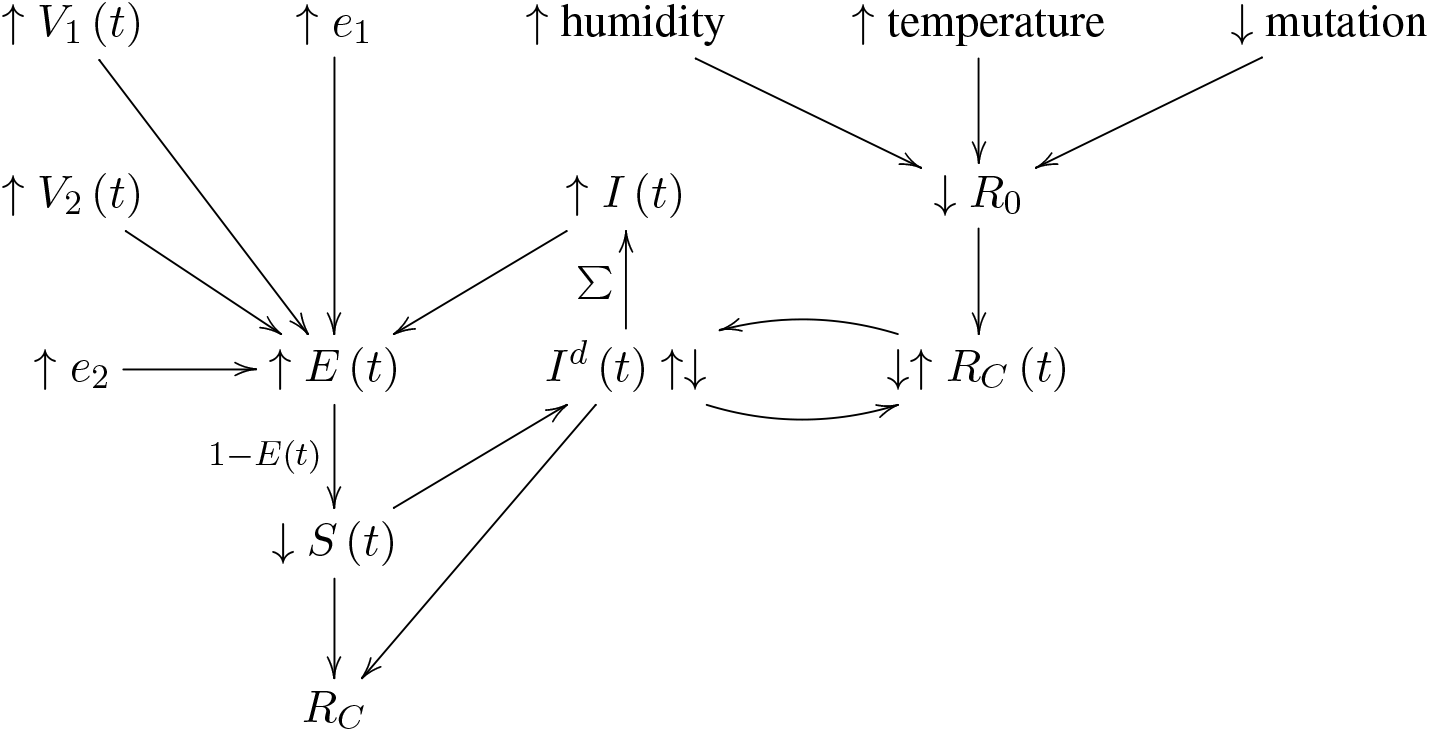

That is, the intrinsic ℛ_0_ can be altered as the humidity, temperature, and mutate variation of the virus changes. Based on the intrinsic ℛ_0_, ℛ_*c*_ (*t*) ≤ ℛ_0_ can be derived which changes through time. That is, the community as a whole follows cycles of relaxation and restrictive measures depends on the observed daily infection. When the infection numbers *I*^*d*^ (*t*) are high, the observant community resorts to more cautious behavior. Which in turn reduces ℛ_*c*_ (*t*) and lower *I*^*d*^ (*t*) is observed. ℛ_*c*_ (*t*) is not the only factor influences *I*^*d*^ (*t*). The remaining susceptible population ratio *S* (*t*) also plays a role in shaping the final numbers of *I*^*d*^ (*t*). The the summation of *I*^*d*^ (*t*) leads to cumulative number of infection *I* (*t*). At the same time, the daily vaccination number *V*_1_ (*t*), the efficacy *e*_1_, the daily vaccination number *V*_2_ (*t*), the efficacy *e*_2_, and cumulative number of infection *I* (*t*) leads to immunity ratio *E* (*t*), which leads to the remaining susceptible population ratio *S* (*t*) = 1 − *E* (*t*). Finally, ℛ_*c*_ at the current time can be derived based on the rate of change in *I*^*d*^ (*t*) and *S* (*t*) leads computed, this is how we arrived at the conclusion that COVID-19 has at least a daily ℛ_0_ ≥ 2.2.

## 5 Conclusions

Israel currently observed ℛ_0_ = 2.2 is a very encouraging value, it implies that the pandemic can possibly terminate sooner than most expect [41] [21]. However, very few countries have reached the level of effective immunization to corroborate the validity of this result other than Seychelles. Seychelles has a much lower population at 100,000 but considerable infections at the current time so that its ℛ_*c*_ (*t*) can be measured. However, ℛ_*c*_ (*t*) of Seychelles has so far not yet plateau and ℛ_*c*_ (*t*) > 2.2. Many explanations can be given, illustrated as a table below.

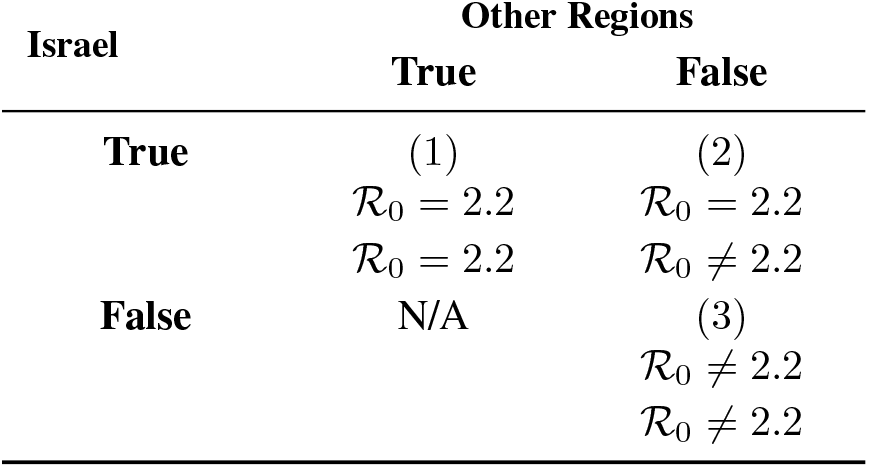

| Israel | Other Regions |  |
| --- | --- | --- |
|  | True | False |
| True | (1)<br>$\mathcal{R}_0 = 2.2$ | (2)<br>$\mathcal{R}_0 = 2.2$ |
| | $\mathcal{R}_0 = 2.2$ | $\mathcal{R}_0 \neq 2.2$ |
| False | N/A | (3)<br>$\mathcal{R}_0 \neq 2.2$ |
| | | $\mathcal{R}_0 \neq 2.2$ |

It is possible that ℛ_0_ = 2.2 is indigenous to Israel only as case (2). Seychelles has a higher ℛ_0_ due to locally higher level of social interaction and different viral strains. Since Seychelles has received a mix dosage of vaccines not limited Pfizer, it is possible that the difference in ℛ_0_ is due to differential efficacy of the vaccines. It is also possible that ℛ_0_ > 2.2. The currently observed value in Israel is still not the intrinsic ℛ_0_ due to continued partial social distancing measures in place as suggested under (3). More analysis is required in the future as more countries reaching full effective immunization ratio. The ℛ_0_ calculation can be replaced by the matrix tabulation we mentioned prior to gain better precision. However, it does not undermine our current assessment, as long as one adopt the identical methodology on computing ℛ_0_ across all data. Finally, even assuming ℛ_0_ > 2.2, due to aggressive vaccination program, continued implementation of restrictive measures, or both, in all countries we have analyzed present an optimistic outlook at controlling the pandemic toward the latter part of 2021.

## Data Availability

Data are available [Edouard Mathieu/ at https://github.com/edomt] with the permission of [Github]. The data that support the findings of this study are publicly available from the corresponding author, [Edouard Mathieu].
Data are available [Worldometer/ at https://www.worldometers.info/] with the permission of [Worldometer]. The data that support the findings of this study are publicly available from the corresponding author [Worldometer]

https://github.com/owid/covid-19-data/blob/master/public/data/vaccinations/country_data/United%20States.csv

https://github.com/owid/covid-19-data/blob/master/public/data/vaccinations/country_data/Israel.csv

https://github.com/owid/covid-19-data/blob/master/public/data/vaccinations/country_data/United%20Kingdom.csv

https://github.com/owid/covid-19-data/blob/master/public/data/vaccinations/country_data/Serbia.csv

https://www.worldometers.info/coronavirus/country/uk/

https://www.worldometers.info/coronavirus/country/israel/

https://www.worldometers.info/coronavirus/country/serbia/

https://www.worldometers.info/coronavirus/country/us/

